# Quantitative EEG Spectral Features Differentiate Genetic Epilepsies and Predict Neurologic Outcomes

**DOI:** 10.1101/2024.10.09.24315105

**Authors:** Peter D. Galer, Jillian L. McKee, Sarah M. Ruggiero, Michael C. Kaufman, Ian McSalley, Shiva Ganesan, William K.S. Ojemann, Alexander K. Gonzalez, Quy Cao, Brian Litt, Ingo Helbig, Erin C. Conrad

## Abstract

EEG plays an integral part in the diagnosis and management of children with genetic epilepsies. Nevertheless, how quantitative EEG features differ between genetic epilepsies and neurological outcomes remains largely unknown. Here, we aimed to identify quantitative EEG biomarkers in children with epilepsy and a genetic diagnosis in *STXBP1*, *SCN1A*, or *SYNGAP1*, and to assess how quantitative EEG features associate with neurological outcomes in genetic epilepsies more broadly.

We analyzed individuals with pathogenic variants in *STXBP1* (95 EEGs, *n*=20), *SCN1A* (154 EEGs, *n*=68), and *SYNGAP1* (46 EEGs, *n*=21) and a control cohort of individuals without epilepsy or known cerebral disease (847 EEGs, *n*=806). After removing artifacts and epochs with excess noise or altered state from EEGs, we extracted spectral features. We validated our preprocessing pipeline by comparing automatically-detected posterior dominant rhythm (PDR) to annotations from clinical EEG reports. Next, as a coarse measure of pathological slowing, we compared the alpha-delta bandpower ratio between controls and the different genetic epilepsies. We then trained random forest models to predict a diagnosis of *STXBP1*, *SCN1A*, and *SYNGAP1*. Finally, to understand how EEG features vary with neurological outcomes, we trained random forest models to predict seizure frequency and motor function.

There was strong agreement between the automatically-calculated PDR and clinical EEG reports (*R*^2^=0.75). Individuals with *STXBP1*-related epilepsy have a significantly lower alpha-delta ratio than controls (*P<*0.001) across all age groups. Additionally, individuals with a missense variant in *STXBP1* have a significantly lower alpha-delta ratio than those with a protein-truncating variant in toddlers (*P<*0.001), children (*P*=0.02), and adults (*P<*0.001). Models accurately predicted a diagnosis of *STXBP1* (AUC=0.91), *SYNGAP1* (AUC=0.82), and *SCN1A* (AUC=0.86) against controls and from each other in a three-class model (accuracy=0.74). From these models, we isolated highly correlated biomarkers for these respective genetic disorders, including alpha-theta ratio in frontal, occipital, and parietal electrodes with *STXBP1*, *SYNGAP1*, and *SCN1A*, respectively. Models were unable to predict seizure frequency (AUC=0.53). Random forest models predicted motor scores significantly better than age-based null models (*P<*0.001), suggesting spectral features contain information pertinent to gross motor function.

In summary, we demonstrate that *STXBP1*-, *SYNGAP1-*, and *SCN1A*-related epilepsies have distinct quantitative EEG signatures. Furthermore, EEG spectral features are predictive of some functional outcome measures in patients with genetic epilepsies. Large-scale retrospective quantitative analysis of clinical EEG has the potential to discover novel biomarkers and to quantify and track individuals’ disease progression across development.

## Introduction

EEG is an integral part of routine, standard care in children with a known or presumed genetic epilepsy. EEG is typically analyzed visually through manual review and annotation by clinicians. This qualitative process is imperfect, with both low inter-and intra-rater reliability, leading to worse outcomes. For example, individuals without epilepsy have a 20-30% chance of misdiagnosis of epilepsy, and there is 50% interrater reliability in identifying epileptiform discharges from EEG.^1–4^ Furthermore, visual review of EEG limits our sensitivity to detect subtle alterations that may serve as biomarkers of epilepsy diagnoses or disease severity.

Many studies have demonstrated the potential of quantitative biomarkers extracted from both clinical and research EEG to identify, predict, and quantify severity in neurological and psychiatric conditions such as autism spectrum disorder (ASD), attention deficit/hyperactive disorder (ADHD), and Alzheimer’s disease.^5–8^ While quantitative analysis of intercranial EEG (iEEG) in presurgical epilepsy patients has been a prominent segment of epilepsy research for over a decade, scalp EEG has received considerably less attention.^9^ This is in part due to the fact that scalp EEG is significantly more prone to artifacts and noise, resulting in a lower signal to noise ratio. More recent, modern signal processing techniques however can quickly and effectively remove artifacts, and machine learning approaches can identify underlying signals from large samples of noisy data.^10,11^

Identifying quantitative biomarkers of disease severity would be particularly impactful in genetic epilepsies. Many genetic epilepsies, particularly developmental epileptic encephalopathies (DEE), are devastating lifelong diagnoses resulting in seizures as well as impairments in cognition, motor, speech, and social skills.^12,13^ Many clinical trials to treat these disorders are underway or in preparation. However, quantifying the severity and tracking the progression of these disorders is often difficult or unfeasible with traditional methods.^14,15^ Furthermore, many of the tests are long, labor intensive, and require highly specialized training to perform. Finding electrophysiological correlates to these comorbidities and their severities could aid in diagnostics and inform the response to treatment to existing and novel therapies.

Here, we hypothesized that quantitative EEG features differ between genetic epilepsies and correlate with functional outcomes. To test this hypothesis, we developed a pipeline to extract quantitative spectral features from a large dataset of pediatric clinical scalp EEG. We validated our pipeline by comparing automatically-measured posterior dominant rhythm (PDR) against clinical annotations. We then compared quantitative spectral features between patients with genetic epilepsy diagnoses and patients without epilepsy or cerebral disease. Finally, we developed machine learning models using spectral features to predict seizure frequency and motor development. Our findings suggest that quantitative EEG measures extracted from standard clinical EEG can be reliably analyzed at scale to identify electrophysiological biomarkers for disease populations and some clinically relevant outcome measures, tracking disease progression across development.

## Materials and methods

### Participant recruitment

We conducted a retrospective cohort study of patients treated within the Children’s Hospital of Philadelphia (CHOP) Care Network, consisting of a main hospital and 50 satellite clinics. We studied three separate cohorts of patients, described below: a non-epilepsy control cohort, a gene-specific epilepsy cohort, and a broader cohort of individuals with known or presumed genetic epilepsy.

For all individuals in our *STXBP1*, *SYNGAP1*, *SCN1A*, seizure frequency, and our primary gross generalize motor function assessment (GMFM) cohorts, informed consent for participation in this study was obtained from parents of all probands in agreement with the Declaration of Helsinki. For all individuals in our control cohort and a subset of individuals in our expanded GMFM cohort, the institutional review board (IRB) waved the requirement for consent. All data was collected per protocol with local approval from the IRB.

### Control cohort

We first aimed to identify patients without epilepsy or other cerebral disease which could result in electrographic abnormalities, serving as a control cohort with anticipated normal EEGs. We identified patients seen at CHOP with EEGs reviewed by a clinician and marked as normal. From all individuals with a clinically normal EEG, we extracted every ICD9/10 diagnosis code available up until September 25, 2024. We composed a list with three epileptologists of all potential ICD9/10 codes which could affect cerebral function or result in an abnormal EEG (for list of ICD9/10 codes restrictions, see **Supplementary Table 1**). We then filtered for controls with no history before or after their EEG recording of any abnormal diagnosis codes from our composed list. Individuals had to have at least one years of available medical record data following their EEG.

### Genetic epilepsy cohorts

Our analyses focused on three specific genetic epilepsy disorders: *STXBP1*, *SYNGAP1*, and *SCN1A*. The CHOP epilepsy center specializes in these disorders and thus a greater number of individuals and their EEGs were accessible for a more comprehensive analysis. Furthermore, these disorders differ greatly in presentations and outcomes, making them ideal candidates for investigation into the capabilities of quantitative EEG analysis. We also collected a broader cohort of individuals with a known or presumed genetic epilepsy and an EEG to investigate electrophysiological biomarkers of seizure control and gross motor function.

All individuals from our genetic epilepsy cohorts were consented and enrolled in the Epilepsy Genetics Research Project (EGRP) at CHOP, which has been enrolling individuals with known or presumed genetic epilepsy since 2014. All genetic diagnoses were reviewed in a clinical and research setting and, if necessary, reclassified according to the criteria of the American College of Medical Genetics and Genomics (ACMG).

In relevant analyses, EEG recordings were placed in age bins based on the age of the individual at the time of the recording. Age bins included: infant (0.25-1 year of age), toddler (1-4 years of age), child (4-10 years of age), and adult (10-21 years of age). Bins were determined by balancing trade-offs between typical EEG age bins and limited sample sizes in our gene-specific cohorts. Similarly, EEGs below three months of age were removed based on common practice of EEG analyses when examining infants and limited sampling in this age range.^16^

### EEG collection and preprocessing

We included any clinical EEGs from the above cohorts recorded in the context of a non-urgent visit, which included outpatient <60 minute EEGs, outpatient 4-hour EEGs, inpatient longterm monitoring (LTM) EEGs, and longterm ambulatory home monitoring EEGs (**Supplementary Table 2**). All clinical notes relating to inpatient continuous monitoring in the gene-specific cohorts and all scheduled intensive care unit (ICU) recordings in other cohorts were reviewed to ensure they were not miscoded in the medical record as non-urgent and that the patient was not in an altered state leading up to or at the time of the EEG recording. If deemed otherwise, recordings were removed. For multiday recordings, only the first day was taken.

All EEGs were recorded using a standard 10-20 system of electrode placement with a varying number of additional EMG, EOG, and EKG electrodes. The machine reference was an electrode placed on the mastoid between electrodes ‘Cz’ and ‘Fz’. All scans were cropped to a maximum of the first 4 hours to reduce the potential accumulation of artifacts. To ensure consistent sampling rate across EEG recordings, we downsampled all EEGs to 200Hz. Prior to downsampling, we performed a 60Hz harmonic filter and a 2^nd^ order infinite impulse response (IRR) filter applied at 95Hz for anti-aliasing.

To remove artifacts such as ocular movement, blinks, cardiac noise, and muscle artifact, we applied an automated Independent Component Analysis (ICA) pipeline through the Python EEG/MEG processing package MNE-ICALabel, following filtering criteria similar to past work.^17,18^ All scans then had a 0.5-70Hz 2^nd^ order bandpass filter applied. We then applied a Laplacian montage. We followed previously published methods to identify and remove artifact-heavy segments.^19^ We also manually reviewed EEG annotations from our control and gene-specific cohort and developed a search algorithm to identify and remove segments of EEG indicating sleep, photic stimulation, induced hyperventilation, and seizures based on clinician and technician annotations. Only recordings with at least 15 clean four-second epochs (one minute of data) following all filters were used in subsequent analyses (preprocessing described in detail in **Supplementary material**).

### Spectral feature extraction

To estimate EEG bandpower, we applied Welch’s method to each EEG electrode across all clean epochs. We extracted delta (1-4Hz), theta (4-8Hz), alpha (8-13Hz), and beta (13-30Hz) bandpowers. We then calculated the relative power of these bands and the alpha-delta, beta-delta, alpha-theta, and beta-theta power ratios. The median of these features was found across epochs in each electrode. We then measured the median across all electrodes as well as across spatial subgroups: frontal (defined as electrodes ‘Fp1’, ‘Fp2’, ‘F3’, ‘F4’, ‘F7’, ‘F8’, ‘Fz’), temporal (‘T3’, ‘T4’, ‘T5’, ‘T6’), occipital (‘O1’, ‘O2’), parietal (‘P3’, ‘P4’, ‘Pz’), and central (‘C3’, ‘C4’, ‘Cz’).

### Automated posterior dominant rhythm detection

To validate our automated EEG processing pipeline, we compared an automatically-detected posterior dominant rhythm (PDR) to that annotated in the clinical report. Our automated PDR calculation followed previously described methods, optimizing peak detection parameters to a subset of our data with a resolution of 0.25Hz (see **Supplemental methods**).^20^

We hypothesized that the automatically-detected PDR could more precisely capture biologically-relevant features pertaining to neural development than the clinician-annotated PDR. To test this, we compared the ability of the automatically-detected PDR and the clinician-annotated PDR to predict an individual’s age at the time of the scan. Given that PDR has an approximately logarithmic relationship with age from infancy to adulthood (**Supplemental Fig. 2**), we fit a generalized additive model (GAM) fit using a gamma distribution with a logarithmic link function.

We performed 1000 permutations with an 80/20 training testing split, evaluating mean absolute error (MAE) and root mean squared error (RMSE) each time.

### *STXBP1*, *SYNGAP1*, and *SCN1A* random forest models

We hypothesized that EEG spectral features would differ between patients with genetic epilepsies and non-epilepsy control patients. We used spatial spectral features including relative bandpowers and power ratios to train random forest models to predict if an EEG originated from a control or an individual with a specific genetic disorder. Including age, this resulted in 41 features. We developed models for *STXBP1*, *SYNGAP1*, and *SCN1A*, identifying age-matched controls for each individual for a 1:1 target-control ratio (see **Supplemental methods**). To test to what extent EEG changes in the genetic epilepsies were diffuse or focal, we also compared our models to those that contained no spatial information. In other words, spectral features were averaged across all electrodes rather than separately by region, resulting in 9 features including age. Performance was evaluated using leave-one-out cross validation (LOOCV).

An additional three-class random forest model was trained to distinguish a diagnosis of *STXBP1*, *SYNGAP1*, and *SCN1A* from each other. To correct for data imbalance, we performed 1000 bootstraps with an 80/20 training-testing split, resampling each time from *STXBP1* and *SCN1A* cohorts to match the size of the smallest cohort, *SYNGAP1*. Similar to past work, feature importance from all models was determined by calculating the Gini index across LOOCVs.^21^

### EEG models to predict seizure frequency and motor development

We also hypothesized that spectral features differ according to clinical outcomes in individuals with a known or presumed genetic epilepsy. As an estimate of seizure outcomes, we obtained seizure frequencies from outpatient clinical notes. Seizure frequency annotations in CHOP notes are binned into the standardized categories: ‘>1 per day’, ‘>1 per week’, ‘>1 per month’, ‘>1 per 6 months’, >1 per 1 year’, ‘1-2 years ago’, and ‘>2 years ago’. To time-align seizure frequencies to EEGs, we required respective clinical notes to have an accompanying non-urgent EEG between 1 month prior to two weeks after the note, reasoning that seizure frequency can change rapidly and medication changes are more likely following a visit. To simplify our model, we split seizure frequency categories into two categories, “severe” (poor seizure control) consisting of the categories ‘>1 per day’ and ‘>1 per week’ and “moderate” (moderate seizure control) consisting of all other categories. We tested other combinations in our **Supplementary analyses**. Here, we also employed age-matching resulting in a 1:1 “severe” to “moderate” outcome ratio followed by LOOCV.

As an estimate of functional outcomes, we obtained clinically-documented gross motor function measure (GMFM) scores from individuals with a known or presumed genetic epilepsy. GMFM scores are a standard diagnostic test to quantify and track motor function in children.^22–24^ We utilized only GMFM scores with an accompanying EEG collected close in time, scaling for age as motor development is non-linear (see **Supplementary Table 3**). Like our PDR analysis, we trained random forest models with spatial spectral features and null models with just age as a feature, performing 1,000 bootstraps with an 80/20 training-testing split, evaluating MAE and RMSE each bootstrap. To test if our findings were specific to genetic epilepsies, we performed the same analyses on a broader cohort of any individual with a clinical EEG and accompanying GMFM under the same constraints.

### Statistical analysis

Nonparametric multi-group comparisons were assessed with a Kruskal Wallis followed by a Dunn test with a Benjamini-Hochberg correction for multiple testing. Statistical difference across childhood between PTV and missense alpha-delta ratios in individuals with a *STXBP1* variant was evaluated by fitting two generalized additive mixed models (GAMM) with individuals as random intercepts, but one with alpha-delta ratio as a fixed effect and one without it. Model comparison was performed via the R function KRmodcomp which approximates an F-test based on Kenward-Roger.^25^ Mann-Whitney U tests determined differences between variants within age groups. Performance of random forest models was determined primarily through receive operator curves (ROC) and area under the curve (AUC). Statistical difference between ROCs was determined with a DeLong’s test. Similar to previous work, when evaluating performance of models with scalar outcomes (PDR and GMFM) against a null model, we calculated the difference in RMSE and MAE between models each bootstrap, then performed a one-sided t-test comparing RMSE and MAE differences against 0.^26^ In scatterplots, a 95% confidence interval was calculated using the R function geom_smooth() default locally estimated scatterplot smoothing (LOESS) regression model, where standard error of the fit is used to estimate the interval around predicted values. All statistical analyses were performed using the R Statistical Framework.^27^

### Data availability

De-identified power spectral data, outcome measures, and code for primary analyses are available at: https://github.com/galerp/scalp_EEG_DEE. Raw EEG data in a de-identified format will be made available by request to the corresponding author.

## Results

### Real-world EEG can be retrieved from the medical record for comprehensive analysis

Clinical EEG is an integral part of routine clinical care for children with known or suspected epilepsy and other neurological disorders. Nevertheless, clinical EEG data is often difficult to extract in large numbers from hospital data warehouses and convert to a readable file format due to proprietary software. We developed an automated pipeline to overcome these hurdles, retrieving a total of 1957 EEGs from 1585 individuals ages 0.03-38.68 years (median 7.23 years, IQR 2.61-12.37 years). The focus of this study was on three genetic epilepsy disorders: *STXBP1*, *SCN1A*, and *SYNGAP1*. We extracted 95 EEGs from 20 individuals with *STXBP1* ages 0.16-17.77 years (median 2.36 years), 154 EEGs from 68 individuals with *SCN1A* ages 0.30-24.62 years (median 4.35 years), and 46 EEGs from 21 individuals with *SYNGAP1* ages 1.19-27.47 years (median 4.07 years; **Table 1**). As seen in **Fig. 1A**, there were some marked differences in age of recordings between the populations particularly at younger ages, validating ages of seizure onset expected in these disorders.^21,28–30^

**Figure 1.**
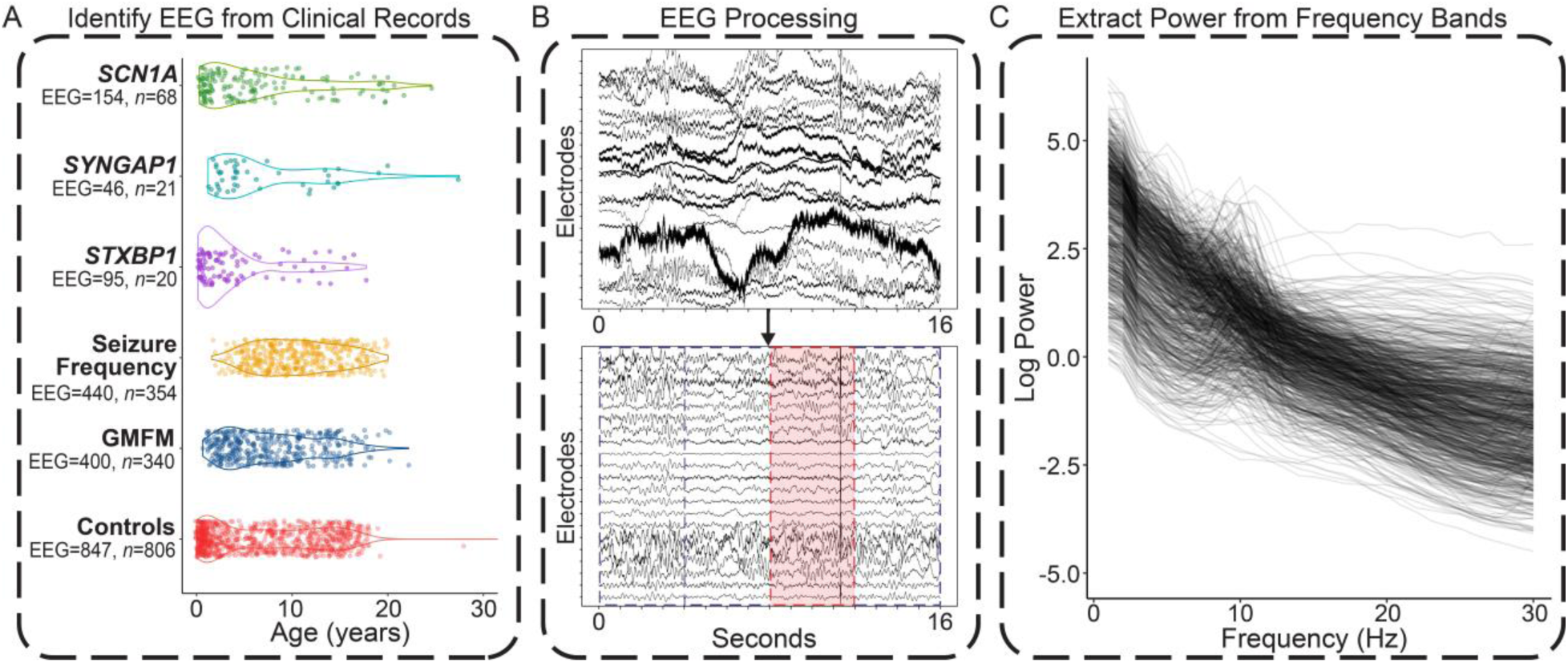
Quantitative EEG pipeline. (**A**) Distribution of EEG recordings in each cohort. Each dot represents a single recording. Two control recordings greater than 30 years of age were removed for figure clarity. (**B**) An example 16 second segment of EEG from an infant with *STXBP1* before and after filters, noise reduction, and artifact rejection. In the cleaned segment, the blue dashed lines indicate clean four-second segments used in downstream analyses. The red dashed lines and red shaded region indicates a segment tagged for removal due to excess artifact, in this case, one or more electrodes exceeding an amplitude of 500µV in the fourth second of that epoch. (**C**) The resulting power spectral densities from all cleaned EEGs from the control cohort recorded after 0.25 years of age. Each line indicates a single EEG recording.

**Table 1:**
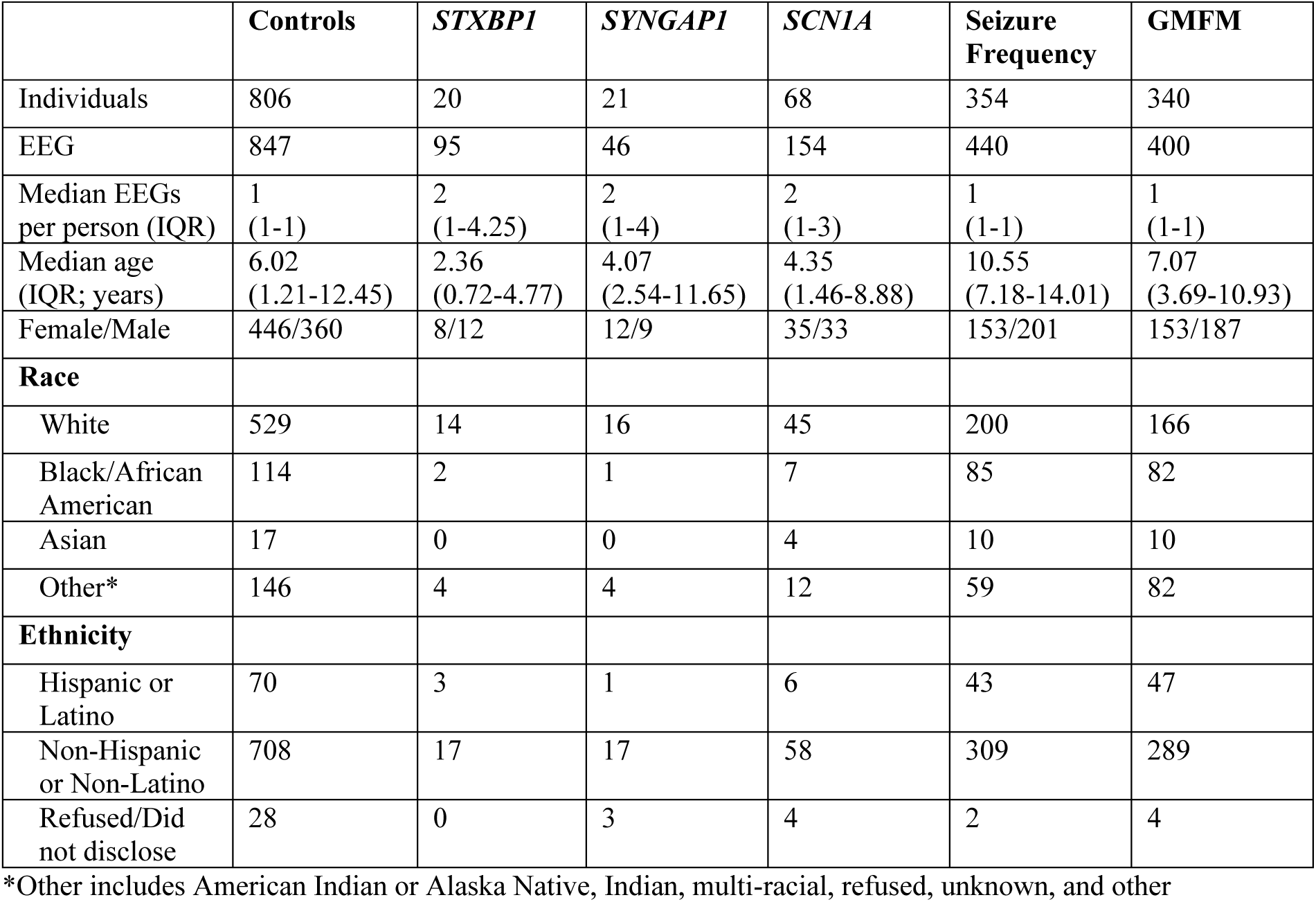
Demographics. Age is calculated across all EEGs thus individuals can be counted multiple times. Race is calculated across individuals thus each individual is counted only once. Seizure frequency represents the cohort of individuals with a known or presumed genetic epilepsy and a seizure frequency clinical annotation with an EEG close in time. Here, the gross motor function measure (GMFM) cohort consists of individuals from the broader analysis of those with and without epilepsy and a GMFM with an accompanying EEG close in time.

A significant obstacle to isolating biomarkers that correlate with neurological abnormalities is the identification of a large cohort of age-matched healthy controls. Leveraging the EMR, we implemented a strict set of criteria to identity a control cohort consisting of 847 EEGs from 806 individuals ages 0.03-38.68 years (median 6.02 years), densely sampling ages across childhood and with a median of 3.98 years (IQR 2.92-5.05 years) follow-up in the medical record after each EEG. We also obtained EEGs from two additional cohorts with standardized clinical measures: 400 EEG (range 0.64-22.18 years of age at recording, median 7.07 years) from 340 individuals with GMFM scores and 440 EEG (range 1.59-20.03 years of age at recording, median 10.33 years) from 354 individuals with a known or presumed genetic epilepsy and a seizure frequency annotation (**Table 1**). Our primary analysis of the GMFM cohort utilized 77 individuals with a clinically known or presumed genetic epilepsy, consisting of 91 EEGs (range 1.04-20.04 years of age at recording, median 4.08 years).

### QEEG can provide higher precision than clinical visual review in detecting electrographic correlates of brain maturation

To assess the accuracy of our pipeline, we tested it against known patterns of spectral features and EEG biomarkers that develop across childhood. We see clear differences in the power spectral density across age groups (**Fig. 1C** ungrouped; **Fig. 2A** grouped). When focusing on the occipital electrodes, we see the development of the posterior dominant rhythm (PDR) as individuals age into adulthood as demonstrated by the small bump in the alpha band (**Fig. 2A**).

**Figure 2.**
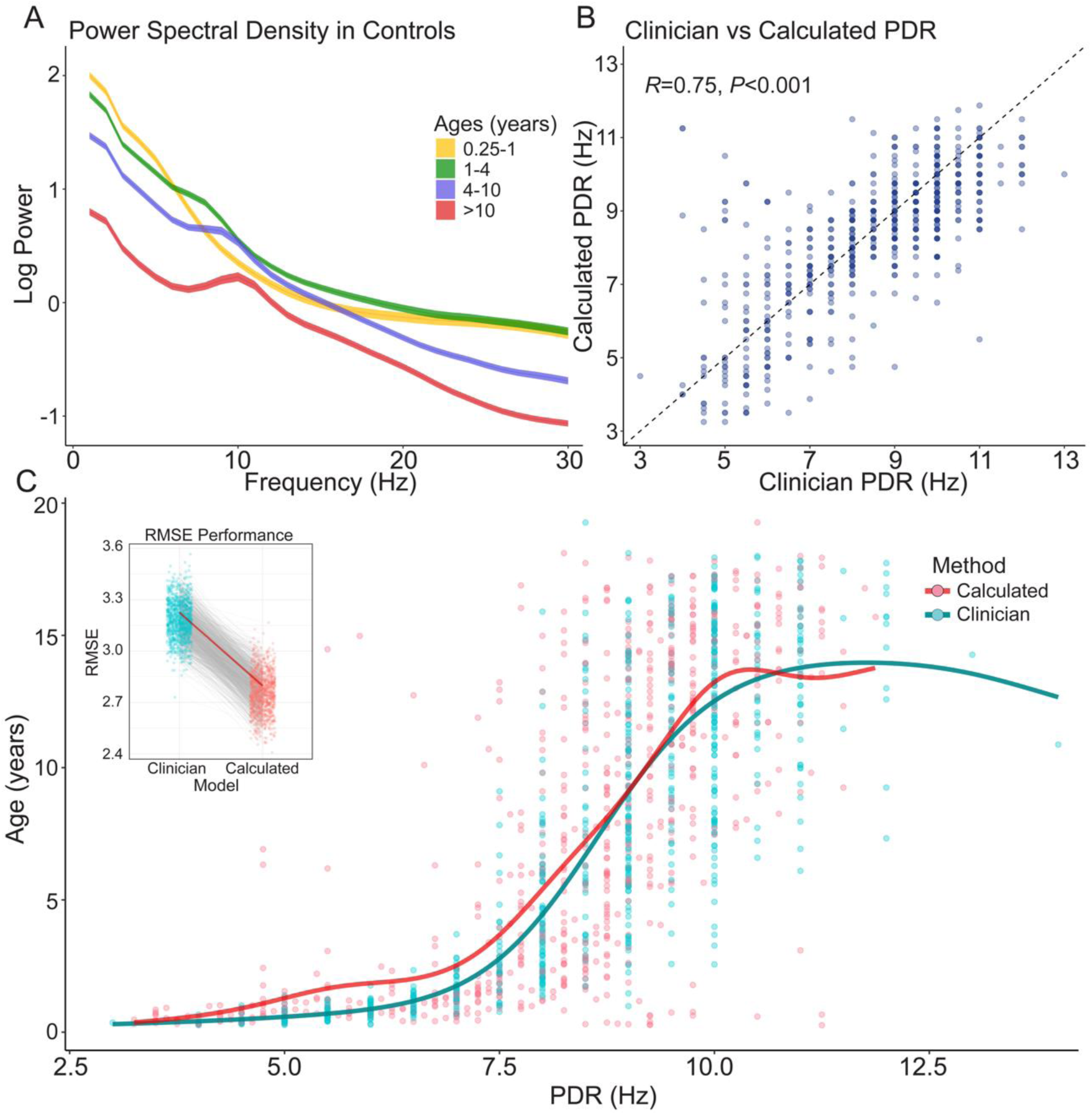
Validation of quantitative EEG pipeline. (**A**) Power spectral density from the occipital electrodes of all EEGs from controls ages 0.25 years and older. Colors indicate age group. Smoothed lines and shading display approximated 95% confidence intervals from a locally weighted smoothing function. Subsequent figures (**B**) and (**C**) display results from analyses comparing calculated vs clinician posterior dominant rhythm (PDR) annotations. (**B**) The correlation between clinician and automated PDR. Each dot is from a single EEG recording. The dotted line is a reference with a slope of one and intercept of zero. (**C**) Automated and clinically annotated PDR across ages. Every EEG from (**B**) is represented as two dots in (**C**), one PDR from clinical annotations (turquoise) and the other from automated extraction (red). The red and turquoise lines indicate a generalized additive model (GAM) fit with a gamma distribution with a logarithmic link function on an 80% sample of the calculated and the clinician PDR respectively. The inset indicates the root mean squared error (RMSE) between the predicted age and actual age across 1000 bootstraps from the clinician model and the calculated model. The grey lines connect the RMSE of each model in the same bootstrap. The red line indicates the permutation with the median difference in RMSE between the two models across permutations.

To further validate our pipeline, we retrieved clinical annotations from 781 EEG reports from respective control EEG from 746 individuals greater than 3 months of age. We could automatically identify a PDR in 737 EEGs (711 individuals; range 0.26-19.27 years, median=7.30 years) with an accompanying clinician annotated PDR. We could not identify a PDR in 31 EEGs with our pipeline. We found that our annotations were highly correlated with those of clinicians (*R^2^*=0.75, *P<*0.001; **Fig. 2B**). While clinician annotated PDR typically has a resolution between 0.5-1Hz, our pipeline had a resolution of 0.25Hz demonstrating the potential of small resolution enhancement with minimal computational cost.

Next we examined whether our extracted PDR provided any additional biologically relevant information beyond what can be gleaned from visual review. We trained and tested logarithmic GAM models to predict age from clinician annotated and our automated PDR (**Fig. 2C**). We found that our models consistently outperformed models trained from clinician annotations, having both a lower mean absolute error (MAE; median=2.38 years) than clinicians (median=2.76 years; *P*<0.001) and lower root mean squared error (RMSE; median=2.76 years) than clinicians (median=3.18; *P*<0.001) across permutations (**Fig. 2C** inset). Taken together, these results suggest that our automated pipeline has high agreement with clinicians for common EEG features and that it may exceed clinician precision at estimating electrographic correlates of brain maturation.

### Alpha-delta ratio correlates with severity in some genetic epilepsy disorders

The alpha-delta ratio is a commonly-investigated quantitative EEG feature in neurological disorders as a rough estimate of the ratio of faster-to-slower activity in the brain that has previously been used to identify sleep.^19,31,32^ We hypothesized that the alpha-delta ratio would be reduced in patients with genetic epilepsies relative to neurologically healthy controls. When examining this feature across age groups and genes in our primary cohort, after multiple testing correction we find that individuals with *STXBP1* exhibit a significantly lower alpha-delta ratio than controls across all age ranges (*P<*0.001, median Cohen’s d=−0.95), *SCN1A* in infancy (*P*<0.001, Cohen’s d=−1.82), childhood (*P*=0.047, Cohen’s d=−0.61), and adulthood (*P*=0.013, Cohen’s d=−1.10) and *SYNGAP1* in toddlers (*P*=3.8x10^-3^, Cohen’s d=−0.68; **Fig. 3A**). Additionally, *SCN1A* demonstrated significantly lower alpha-delta ratio compared to controls in toddlers (*P*=0.013, Cohen’s d=−0.43) and children (*P*=0.042, Cohen’s d=−0.23), and *SYNGAP1* displayed lower alpha-delta ratio than controls in children (*P*=0.033, Cohen’s d=−0.62).

**Figure 3.**
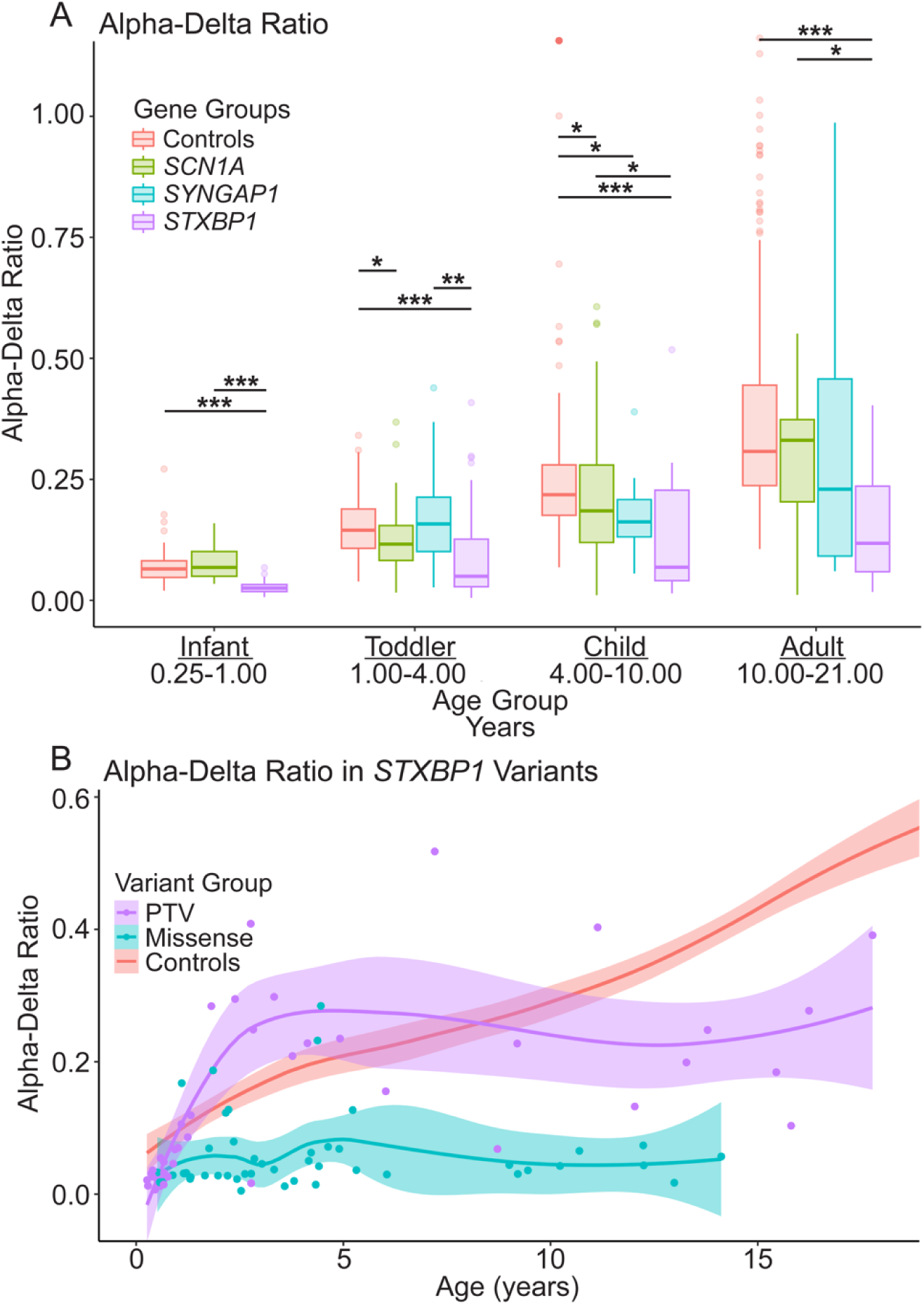
Alpha-delta ratio in *STXBP1*, *SCN1A*, *SYNGAP1*, and Controls across childhood. (**A**) Alpha-delta ratio in discrete age bins (\**P* < 0.05; \*\**P* < 0.01; \*\*\**P* < 0.001). For visualization purposes, 10 data points outside the displayed range were removed (10 adult controls), however, all were included in the analyses and boxplot construction. (**B**) Alpha-delta ratio in individuals with protein-truncating variants (PTV; purple) and missense variants in *STXBP1* (turquoise) and Controls (red). Dots depict a single EEG recording from individuals from one of the *STXBP1* variant groups. Controls displayed for reference. Smooth lines and shaded regions are fitted via locally weighted scatterplot smoothing, the latter approximating a 95% confidence interval.

Alpha-delta ratios also varied within gene groups. We hypothesized that this could be due in part to variant type, which can correlate with disease outcomes. As a secondary analysis, we compared alpha-delta ratios between individuals with missense variants in *STXBP1* to those with protein-truncated variants (PTV; **Fig. 3B**). A generalized additive mixed effect model (GAMM) fit with the inclusion of this variant type variable provided a significantly higher fit to the data than those without it (*P*=0.017).^25^ Examining these results more closely, we find individuals with missense variants in *STXBP1* had a significantly lower alpha-delta ratio than those with PTVs across all age groups except infants (toddler, child, and adult: *P*<0.001, *P*=0.012, *P*<0.001). Overall, these findings support that, relative to neurologically healthy controls, the alpha-delta ratio is reduced in *STXBP1*, and in some age groups in *SCN1A* and *SYNGAP1*.

### Spectral features of EEG can distinguish between *STXBP1*, *SYNGAP1* and *SCN1A* and controls with high accuracy

We next hypothesized that different genetic epilepsies demonstrate distinct patterns of spectral abnormalities that can be probed in a multi-feature machine learning model. We used the median spectral features relative delta, theta, alpha, and beta and the power ratios alpha-delta, beta-delta, alpha-theta, and beta-theta in five pre-defined regions of the scalp. We applied these features along with age of the individual at the time of the EEG to train random forest models to make binary predictions between a particular genetic epilepsy disorder (*STXBP1*, *SYNGAP1*, and *SCN1A*) and age-matched controls.

*STXBP1* random forest models outperformed all other gene prediction models, achieving an AUC of 0.91, F1 of 0.81, precision of 0.86, and recall of 0.76 (**Fig. 4A**). The most informative features originated predominantly from frontal electrodes (**Fig. 4B**). We could predict *SYNGAP1* with an AUC of 0.82 and F1 of 0.71, precision of 0.71, and recall of 0.71 (**Fig. 4C**), and *SCN1A* from controls with an AUC of 0.86, F1 of 0.78, precision of 0.78, and recall of 0.77. The two most important features localized to occipital electrodes in *SYNGAP1* models and in parietal electrodes in *SCN1A* models (top features in **Supplementary Fig. 5-7**; feature correlations in **Supplementary Fig. 8-11**). All non-spatial models displayed a significant drop in performance (*STXBP1 P*<0.001, *SYNGAP1 P*=4.0x10^-3^, *SCN1A P*<0.001) as calculated by the DeLong test, suggesting the importance of spatial information in distinguishing genetic epilepsies.

**Figure 4.**
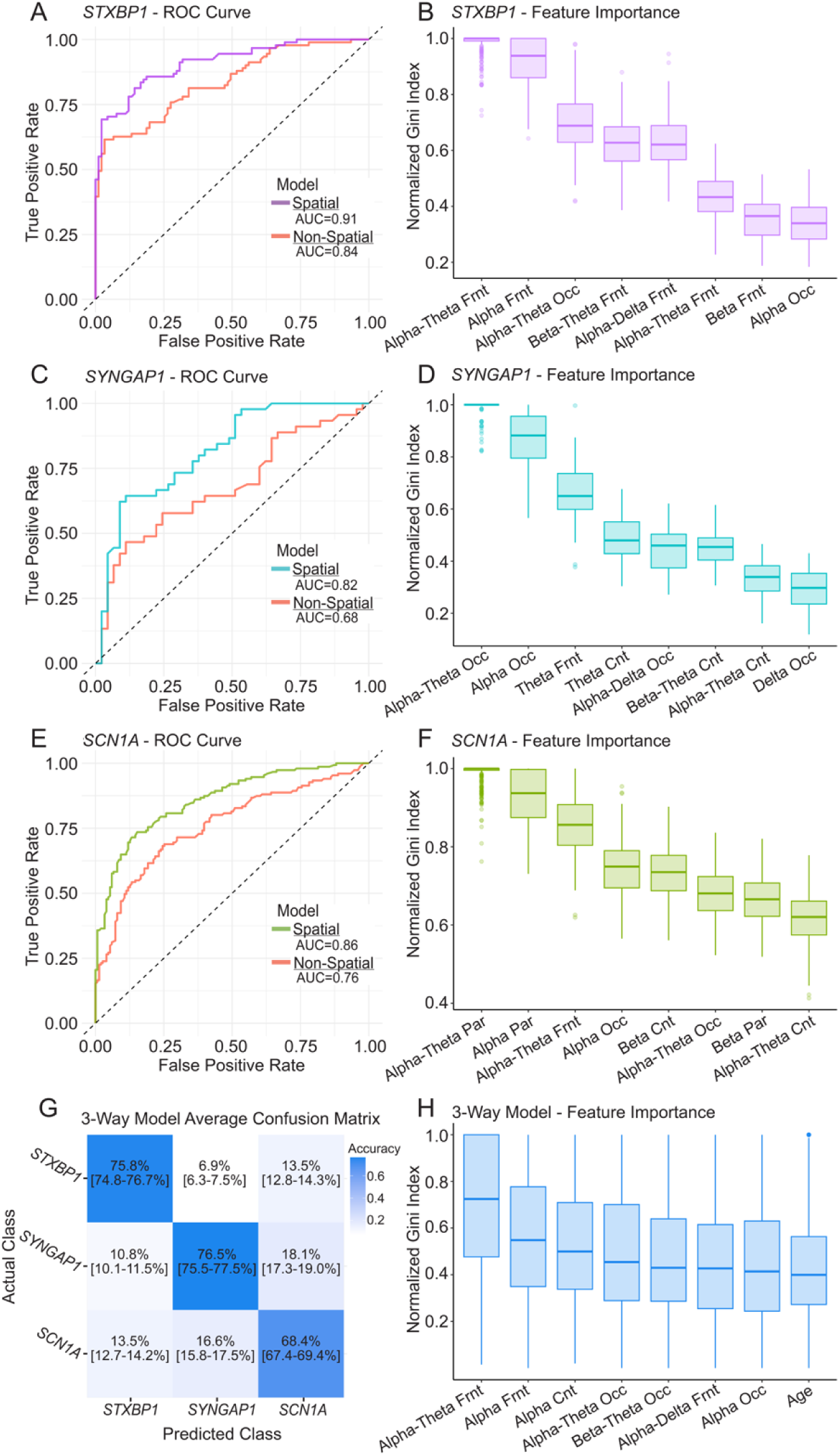
Random forest models to predict a diagnosis of *STXBP1*, *SYNGAP1*, and *SCN1A*. Random forest models trained on spectral features across 1000 bootstraps to predict *STXBP1* [(**A**) and (**B**)], *SYNGAP1* [(**C**) and (**D**)], *SCN1A* [(**E**) and (**F**)] from age-matched controls. ROC curves in (**A**), (**C**), and (**E**) displaying performance with colored lines indicating performance in models trained with localized spectral features, and red lines indicating performance in models with median spectral feature values across all electrodes. The performance of a 3-way model predicting *STXBP1*, *SYNGAP1*, and *SCN1A* from each other is displayed in a confusion matrix in (**G**). Numbers indicate mean percent guessed by the model in that category thus the diagonal indicating percent accuracy in predicting the respective gene. The shading reflects these mean percentage values, with darker blues indicating a higher relative percentage. Numbers in brackets indicate 95% confidence interval. The top 8 most important features as measured by the Gini index normalized across permutations is displayed in the boxplots (**B**), (**D**), (**F**), and (**H**). Features were chosen and ordered by their median normalized Gini index across permutations.

To compare genes to each other, we trained a three-class random forest model. When examining binary performance, one-vs-all, it achieved high performance with *STXBP1* achieving a median AUC of 0.92 (IQR 0.88-0.96), *SYNGAP1* a median AUC of 0.91 (IQR 0.86-0.96), and *SCN1A* a median AUC of 0.88 (IQR 0.82-0.93). Models with just age as a feature had a lower performance: *STXBP1* with a median AUC of 0.73 (IQR 0.65-0.80), *SYNGAP1* a median AUC of 0.82 (IQR 0.76-0.88), and *SCN1A* a median AUC of 0.67 (IQR 0.60-0.76); suggesting age differences between genetic epilepsies do not fully explain model performance. The model achieved strong three-way classification performance with median F1s of 0.78, 0.75, and 0.70 in *STXBP1*, *SYNGAP1*, and *SCN1A* respectively. In summary these models demonstrate that spectral features can accurately distinguish individuals with *STXBP1*, *SCN1A*, and *SYNGAP1* from controls and, to a lesser degree, each other across development. Furthermore, from these models we can extract the ensemble of biomarkers that aid in this discrimination and roughly isolate spatial importance of different brain regions.

### Seizure severity displays no spectral EEG correlates

We hypothesized that spectral features may also contain information correlated with seizure control in individuals with a known or presumed genetic epilepsy. We extracted 440 clinical documentations of seizure frequency collected across 354 individuals each with an accompanying EEG. Five individuals across seven EEGs were removed due to insufficient EEG epochs after filters. Scores were split into the binary categories “severe” and “moderate” (**Fig. 5A**). As age alone provided moderate predictive ability, we performed age matching which also balanced classes. We found models performed no better than chance (AUC=0.53, **Fig. 5B**). While alterations in the binary split of the categories resulted in small boosts in performance (**Supplementary Fig. 1**), overall, these results suggest spectral features do not contain correlates to seizure control.

**Figure 5.**
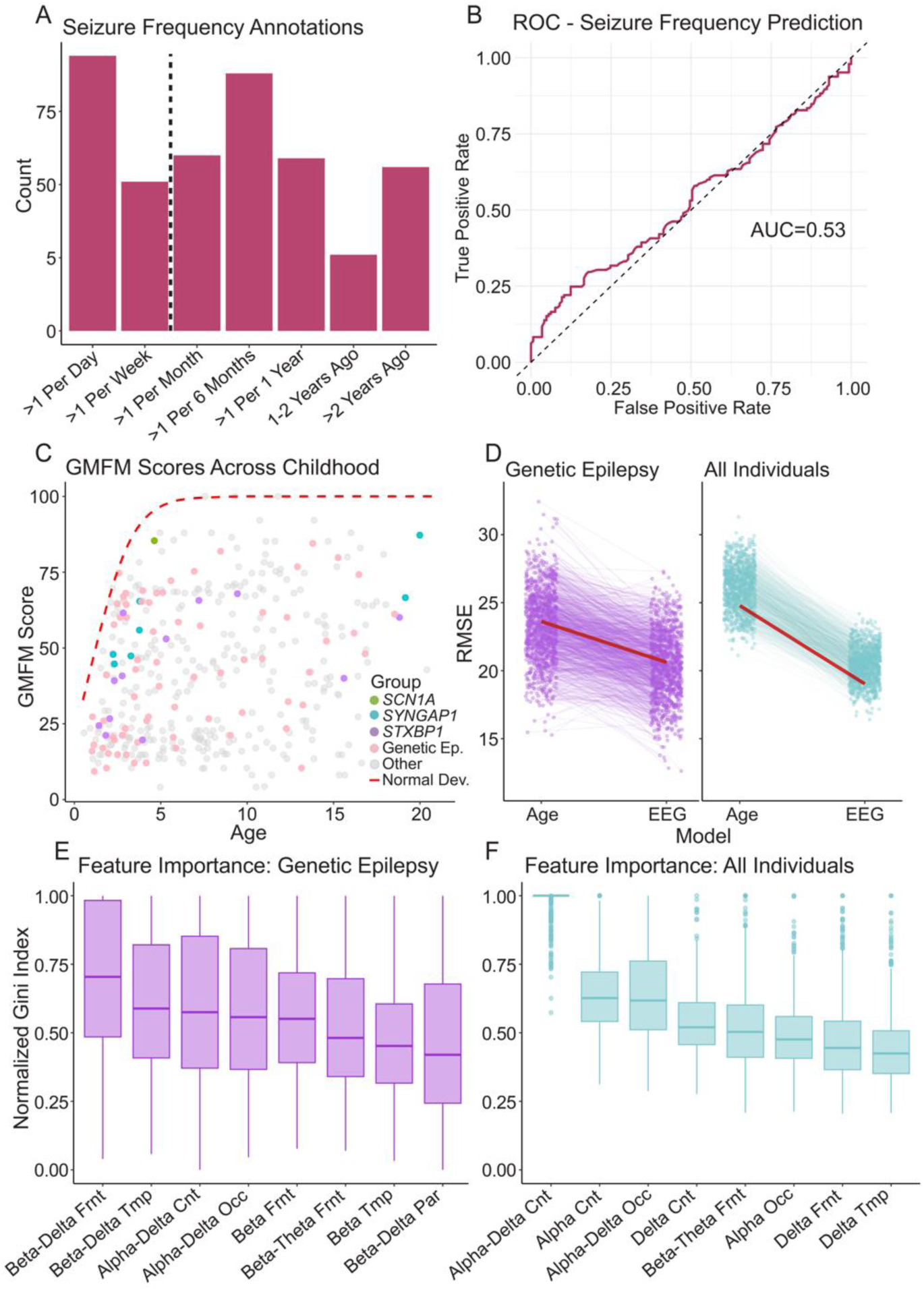
Ability of spectral features to predict outcome measures of seizure frequency and motor development. (**A**) Distribution of seizure frequency annotations from individuals with a known or presumed genetic epilepsy and an accompanying scalp EEG close in time that passed all filters. The dotted vertical line indicates where the data was split into binary outcomes: “severe” and “moderate” seizure control. The performance of the model using age-matching between classes followed by leave-one-out cross validation is displayed in the ROC curve in (**B**). (**C**) All generalized motor function measure (GMFM) scores by age in years with an accompanying scalp EEG close in time. Colors indicate the disease group with light gray representing any individual without a known or presumed genetic epilepsy. This group contains individuals with and without epilepsy. The dotted red line indicates expected GMFM scores for normal development (Schwartz et al., 2021). (**D**) Random forest models’ performance trained to predict a GMFM from individuals with a known or presumed genetic epilepsy on the left (purple) and all individuals on the right (turquoise). Performance was measured by root mean squared error (RMSE). Lines indicate the same bootstrap between the primary and comparator age model. The red line indicates the bootstrap with the median difference in RMSE between each model. Normalized feature importance of the primary models, as measured by Gini index is displayed in (**E**) and (**F**) for models trained on individuals with a known or presumed genetic epilepsy and all individuals respectively. Features and their order were determined by the median normalized Gini index across bootstraps.

### EEG spectral features contain correlates with abnormal motor function

We collected 91 complete GMFM scores (median score=44.7; IQR 23.90-63.00), a standardized measure of gross motor development, from 77 individuals (median=4.19 years of age at time of GMFM, IQR 2.77-9.32 years) with a known or presumed genetic epilepsy. One EEG-GMFM pair was removed due to insufficient epochs in their EEGs after filters. As demonstrated by the red dashed line in **Fig. 5C**, in individuals with typical motor development, GMFM scores progresses exponentially before plateauing and reaching a perfect score of 100 at around 5 years of age.^23,24^

Again, we tested a series of machine learning models using the same set of spatial spectral features along with age at the time of GMFM assessment. We compared these against a null model with only age as a feature. Across bootstraps, random forest models with age and spectral features consistently outperformed null models achieving a significantly lower MAE (*P*<0.001; median EEG model MAE=16.9; median difference=2.6) and lower RMSE (*P*<0.001; median RMSE=20.4; median difference=3.0; **Fig. 5D**).

To test whether these findings were specific to individuals with a genetic epilepsy, we expanded the cohort to any individual with a GMFM and EEG at a similar point in time, resulting in 340 individuals across 400 unique GMFM-EEG data pairs recorded across 0.64 to 22.18 years of age (median=7.10 years, IQR 3.71-10.95 years), removing 6 GMFM-EEG prior to analysis due to excess noise. Results improved relative to null models with the larger, broader cohort in both RMSE (*P*<0.001; median EEG model RMSE=20.4; median difference=5.8; **Fig. 5D**) and MAE (*P*<0.001; median MAE=17.1; median difference=4.3), with age becoming a worse predictor of motor function. We found alpha-delta ratio in the central and occipital electrodes to be consistently among the most important features across both models (**Fig. 5E** and **5F**).

In summary, this consistently significant superior performance suggests that EEG contains some electrographic correlates of motor development, and this phenomenon is not restricted to individuals with a genetic epilepsy.

## Discussion

We analyzed the capability of an automated quantitative scalp EEG pipeline to identify known and novel biomarkers in a heterogenous dataset of 1957 EEGs from 1585 individuals 0.03-38.68 years of age. Scalp EEG is an integral part of routine clinical care in children with a genetic epilepsy. Nevertheless, analysis is largely manual and prone to error. The depth of information held within the electrophysiological recordings remains largely unexplored despite great technological advances, particularly in the fields of AI and machine learning.^11^ Much of the past research of quantitative EEG in childhood disorders often struggles to obtain a sufficient sample size, particularly in control populations. Consequently, the age dimension is often ignored or treated linearly. Here, we demonstrate the ability of an automated quantitative EEG pipeline to identify EEG signatures of *STXBP1*-, *SYNGAP1*-, and *SCN1A*-related epilepsy disorders and to predict neurological outcomes.

### A quantitative EEG processing pipeline identifies expected EEG signatures of normal brain maturation

We demonstrate the ability to leverage the EMR to isolate a control population with normal electrophysiology. While all EEGs were manually reviewed and marked normal by a clinician, there still may be underlying abnormalities. By collecting all available diagnosis codes from controls and requiring at least one year of patient follow-up after the EEG recording, we can minimize the potential of individuals with electrophysiological abnormalities. Supporting this claim, we find the most common ICD10 code in this cohort is “Transient alteration of awareness” (R404; *n*=385) and “Unspecified abnormal involuntary movements” (R259; *n*=271) is the seventh most common, suggesting an EEG was performed to rule-out seizure as the etiology of a transient event or other related abnormality.

To provide further validation of this cohort and our automated methodology, we compared the PDR extracted from our automated pipeline to that gathered through manual review by clinicians. We closely match clinician annotations with an *R^2^* of 0.75 (*P*<0.001). We also found that a model incorporating quantitative PDR estimates predicted the age of the individual with significantly higher accuracy than one incorporating clinician estimates. This may reflect higher precision and resolution in automated PDR estimates and suggests that even minor differences in PDR reflect actual changes in brain maturation. It remains unknown whether subtle deviations in the PDR below the threshold of clinician annotations are associated with neurological disease. Taken together, this provides evidence of the reliability and robustness of our pipeline and the potential benefit such automated techniques have in the clinic for even routine measures such as PDR.

### Alpha-delta ratio differentiates patients with genetic epilepsies from controls

The alpha-delta ratio has been repeatedly implicated as an estimate slowing in the brain and impaired cognition. It has been shown to be statistically different in several neurological conditions such as Alzheimer’s disease, traumatic brain injury outcomes, ischaemic stroke, and *CDKL5* deficiency disorder and an estimate of sleep.^19,31–34^ We hypothesized that alpha-delta ratio may be diminished in certain genetic epilepsy populations, particularly those with more severe developmental comorbidities. Here, we demonstrate that across all age groups individuals with an *STXBP1*-related epilepsy disorder have a significantly lower alpha-delta ratio than controls and some other genetic epilepsy populations at certain ages (**Fig. 3A**). This supports past findings which, utilizing a novel metric, found a lower excitation-inhibition ratio in EEGs of individuals with an *STXBP1*-related disorder.^35^ Furthermore, *STXBP1* tends to present with more severe developmental impairment than *SCN1A*-and *SYNGAP1*-related disorders.^28,36^

We also found that much of the variability in alpha-delta ratio among individuals with *STXBP1* could be explained by their variant. Those with a missense variant in *STXBP1* tend to have a stagnate alpha-delta ratio from infancy onward, while those with a PTV have a ratio which rises similarly to controls up until approximately 4 years of life. This matches what has been observed clinically in individuals with *STXBP1* disorders. It has been repeatedly observed that individuals with recurrent missense variants in *STXBP1* that account for a large proportion of missense variants have a more severe presentation given a presumed dominant-negative effect.^28,36^ As seen in **Fig. 3B**, several data points from three individuals with a PTV do not match this trend, however, this may be in part explained by disease progression (see **Supplemental material**).

Importantly, we see that these changes, along with most spectral features, are largely nonlinear and vary in trajectory and variance within the disease population (**Fig. 3A**; **Supplementary Fig. 3-7**), emphasizing the importance of age and the statistical methods by which these signals must be examined.

### EEGs display distinct electrophysiological backgrounds in *STXBP1*, *SYNGAP1*, and *SCN1A*

We found that EEG spectral features could be used to predict genetic epilepsies from controls with high accuracy, achieving an AUC of 0.91, 0.82 and 0.86 in *STXBP1*, *SYNGAP1*, and *SCN1A* respectively. This high performance is striking considering the known heterogeneity within these disorders’ outcomes and presentations and the wide range of ages sampled.^15,28,29,37^ Additionally, in some of these individuals, clinicians may not observe any abnormal activity in their EEG.^38^ For example, clinicians observe normal EEGs in 43% of infants and 10% of older individuals with *SCN1A*-positive Dravet syndrome.^38^ Secondly, many individuals with a pathogenic variant in *SCN1A* can present with mild phenotypes. In our *SCN1A* cohort, 18 of 68 individuals had typical development as confirmed by manual chart review.

As a preliminary investigation, we tested the ability of a model to distinguish between the three disorders. When examining binary performance, one-vs-all, it achieved high performance with *STXBP1* achieving a median AUC of 0.92 (IQR 0.88-0.96), *SYNGAP1* a median AUC of 0.91 (IQR 0.86-0.96), and *SCN1A* a median AUC of 0.88 (IQR 0.82-0.93; **Fig. 4G**). Our inability to use age matching in this model due to diffuse sampling, however, makes it difficult to compare this performance to our primary models trained with controls. Notably, across all models alpha-theta ratio was the top feature. Repeatedly shown to correlate with cognitive deficits and decline in other diseases, alpha-theta ratio could be a fruitful area of further investigation in the genetic epilepsies.^39–41^ In summary, our findings suggest that individuals with *STXBP1*, *SYNGAP1*, and *SCN1A* have underlying distinct spectral dynamics in their wake EEG that distinguish them from controls and each other.

The higher performance of models that incorporated spatial information relative to spatially agnostic features suggests that EEG abnormalities are to some degree focal in these diseases,^38,42^ which challenges our traditional clinical view of these disorders as diffuse cerebral processes. The specific location of EEG abnormalities generates hypotheses for future studies into the biological mechanisms underlying phenotypes in specific genetic epilepsies. For instance, in *STXBP1* we found alpha-theta, alpha, and beta-theta in the frontal lobe and alpha-theta in the occipital lobe to be the top four most important features. Further carefully orchestrated hypothesis driven experimentation could identify novel biomarkers of disease and severity and endpoints for clinical trials.

### Seizure frequency does not affect underlying spectral dynamics

The primary aim of care for most epilepsy disorders is seizure control. However, self-reported seizure frequency is often unreliable, particularly with individuals that live alone.^43^ Past work has indicated that seizure frequency often correlates with quality of life and severity of other comorbidities.^44–46^ Nevertheless, we found that spectral features were indistinguishable between those with poor and moderate seizure control and a known or presumed genetic epilepsy. Follow-up analyses on a subset of individuals with any genetic epilepsy diagnosis similarly performed no better than chance. These results suggest that seizure control does not significantly impact underlying spectral features in awake individuals with genetic epilepsy. Future directions include testing whether other quantitative EEG biomarkers, such as automated spike rate detections, vary with seizure frequency.^47^

### Spatial spectral features hold information indicative of motor development

Motor developmental delay and abnormalities are a common comorbidity in the childhood epilepsies and other neurodevelopment conditions. We sought to determine if quantitative characteristics of EEG held information pertinent to functional status, in particular, gross motor ability. We extracted 91 gross motor function measures (GMFM) with an accompanying EEG recording from 77 individuals with a known or presumed genetic epilepsy. We found that random forest models trained with spatial spectral features from individuals’ EEG performed significantly better at predicting GMFM than a model trained with just age as a feature (*P*<0.001; **Fig. 5D**). Relative performance improved when including any individuals with a GMFM and EEG. These findings suggest that some EEG spectral changes reflect motor function status. Future models with more complex feature sets may achieve even higher accuracy, suggesting the possibility of using EEG to track functional status in clinical trials and other applications.

### Limitations

There were several notable limitations in our study. First, although we took steps to exclude patients from our control cohort with diagnoses that would suggest cerebral disease, it is possible that some of these individuals are not truly neurologically typical. Importantly, the inclusion of individuals with neurological disease in our control cohort would likely reduce observed differences between controls and our genetic epilepsy cohorts, resulting in reduced power rather than an increased rate of Type I errors. Similarly, there may have been a surveillance bias towards individuals with worse seizure control. Such individuals are more likely to obtain repeat EEGs. However, as demonstrated by the poor performance of our seizure control models, their spectral features were likely minimally affected by this.

Secondly, scalp EEG is inherently noise ridden. Cleaning this signal can be particularly difficult in children with behavioral abnormalities and sensory sensitivities, both of which are common in genetic epilepsies. It is possible that EEG artifacts differ between children with genetic epilepsies and our control cohort, contributing to some of our model performance. To mitigate this concern, we use conservative filters, including an ICA, strict thresholding for epochs, and removal of events as marked by clinician and technician annotations. As a result, we may have removed some biologically relevant signals.

Additionally, annotations alone could not ensure a consistent resting state across all individuals and their recordings. Medication, particularly benzodiazepines, have been also shown to potentially impact EEG and were likely prescribed to some individuals in these cohorts.^48^ These, however, were very unlikely prescribed across an entire cohort. Consequently, their effects are likely insufficient to achieve the accuracies demonstrated by our models.

Finally, in our analyses we used relatively simple spectral features. Past work, particularly in intercranial EEG, has shown the benefit of more complex network features.^9,49,50^ The importance of the spatial dimension in our results suggests the interplay between these regions may hold significance.

## Conclusion

In summary, we developed an automated quantitative EEG pipeline to retrospectively identify and gather nearly 2000 clinical scalp EEGs and extract relevant spectral features. Our results suggest that the genetic epilepsies have distinct quantitative electrographic signatures. We demonstrate that with minimal clinician review, we can identify novel biomarkers for three distinct genetic epilepsy disorders, *STXBP1*, *SYNGAP1*, and *SCN1A*. Furthermore, our analyses suggest underlying electrophysiological activity suggestive of motor development. Additionally, we can track these changes across childhood, further demonstrating the importance of age as a feature when analyzing EEG. The approach and features outlined in this work can contribute to large-scale quantitative EEG analysis, novel biomarker discovery, and more precise tracking of clinical outcomes and severity across development in a diverse set of neurological disorders.

## Data Availability

All deidentified data produced are available online at https://github.com/galerp/scalp_EEG_DEE/.

## Acknowledgments

We thank the participants and their family members for taking part in the study. We would like to thank Mahgenn Cosico and Priya Vaidiswaran for their support in enrolling research participants and for administrative assistance. We would also like to thank Sam Pierce, PT, PhD for his advising on the GMFM.

