## Supplementary material for "Quantitative EEG Spectral Features Differentiate Genetic Epilepsies and Predict Neurologic Outcomes"

#### EEG preprocessing

Our EEG processing begins with ensuring that the recording is no longer than four hours in length. If the file is found to be longer than this, it is cropped to the first four hours of the recording. Then the sampling rate is checked. If the sampling rate exceeds 200Hz (the majority of the recordings), the recording is downsampled to 200Hz. Prior to downsampling, however, a 60Hz harmonic firwin notch filter is applied to remove electrical noise, then a lowpass infinite impulse response (IIR) filter is applied at 95Hz (5Hz less than the Nyquist frequency). The signal is then converted from microvolts to volts.

A small subset of EEG from 2023 and 2024 used the updated Modified Combinatorial Nomenclature (MCN) which utilizes additional electrodes as well as renaming four electrodes present in the original 10-20 system. If these new channel names are detected, the renamed four electrodes are converted back to their original names. To remove artifacts such as those arising from eye movement, blinks, heartbeat, and muscle, we then applied an automated Independent Component Analysis (ICA) pipeline through the Python EEG/MEG processing package MNE-ICALabel. We followed past work and utilized settings as close as possible to those suggested by the original package designers.[1,2] In short, all potential channels seen in the EEGs in our cohorts were set to their applicable channel types (“eeg”, “eog”, “emg”, “ecg”, “misc”). This aids the algorithm in identifying artifacts in the recordings. Channels with patient events were removed (e.g., “photic stimulation”) prior to this filtering step. A 2$^{nd}$ order IRR bandpass filter is applied between 0.5-95Hz. The ICA function is then applied, extracting 20 components. Components are given a set of probabilities attached to labels including ‘brain’, ‘muscle artifact’, ‘eye blink’, ‘heartbeat’, ‘line noise’, ‘channel noise’, and ‘other’. To be conservative, if a component does not include any likelihood of ‘brain’ or a less than 30% likelihood of ‘other’, it is removed from the signal.

A 0.5-70Hz 2nd order bandpass filter is then applied to the resulting signal, and we then apply a Laplacian montage. Next the signal is placed into non-overlapping one-second epochs. Epochs are then removed if any electrode has a line length or root-mean-squared amplitude greater or equal to two standard deviations above the mean of that electrode or exceeds an absolute amplitude of 500μV. We then apply a custom function to identify epochs that lie near or within segments annotated by EEG technicians or clinicians as containing sleep, seizures, photic stimulation, or induced hyperventilation.

In brief, we examined over 1000 EEGs’ annotations to identify the different spellings and phrases to indicate sleep and seizures. Photic stimulation and hyperventilation are standardized annotations within the hospital system’s software. In the case of photic stimulation, epochs are removed from the start to 20 seconds after the marked end of the stimulation. For hyperventilation, we remove the start to 120 seconds after the marked end. When sleep is annotated (text variations of “sleep”), we take the beginning to the end (marked by some form of “awake”) with no padding. Through our preliminary investigation into a large subset of annotations, we took into account variations of strings that included negation such as “not asleep”. If none of the above annotations has a clearly marked end, the remainder of the recording is removed. When a seizure is annotated, we remove 60 seconds prior to the first annotation and the remainder of the recording.

In the final step, consecutive non-overlapping four-second epochs are extracted, combining the previously demarcated one-second epochs. This means if the final second of a four-second epoch was marked for removal from a previous preprocessing step, the entire epoch would be removed.

For example, in the cleaned EEG segment in **Fig. 1B** the third epoch is removed due to the fourth second in that epoch containing at least one electrode with a signal exceeding 500μV. Consequently, the next four-second epoch begins. EEG recordings are only used in downstream analyses if they have at least 15 four-second epochs (one minute of data) remaining after cleaning.

#### Spectral feature extraction

Using Welch's method with a 1Hz step size, the power spectral density (PSD) is extracted from each epoch of each electrode from the cleaned EEG. We then find the median PSD for each electrode across all available epochs. After extracting the median PSD, the frequencies are split into delta (1-4Hz), theta (4-8Hz), alpha (8-13Hz), and beta (13-30Hz) power bands. This is what is also used for the powerband ratios (e.g., alpha-delta, beta-delta, etc.). To find the relative power of each frequency band, each respective powerband is divided by the sum of the powers of the frequencies 1-70Hz.

#### Posterior dominant rhythm (PDR) detection

Since January 2018, all clinical EEGs at CHOP must be reviewed by a clinician who annotates an accompanying smartform with common data elements based on ACNS (American Clinical Neurophysiology Society) standardized terminology. We collected all smartforms from controls and extracted posterior dominant rhythm (PDR) annotations. If annotations contained a range (e.g., "7-8Hz"), the median was taken. Based on previously published methodology, we developed an automated PDR detector.[3]

Similar to this past published method, a detector took clean 4-second epochs from occipital electrodes (O1 and O2), converted the signal of decibels, applied a Savgold smoothing filter, and then extracted the power spectral density curve with a resolution of 0.25Hz. From each epoch, we isolated the peak frequency and its key features including, prominence, power, width height, and width from each occipital electrode ('O1' and 'O2'). Taking a subset of a random 180 EEG recordings, we manually identified the optimal filters from these features to identify epochs with a PDR, using the raw power spectral density and age of the individual as a guide. These filters include a peak with a frequency of ≤14Hz and ≥3Hz, prominence >5.5, power >6 and <21, width <20, and several others that can be seen in our GitHub repository. We defined PDR as the median peak frequency across the remaining epochs and electrodes.

#### Alpha-delta ratio in *STXBP1* variants

In our primary results, we find that, overall, individuals with a protein-truncating variant (PTV) in *STXBP1* have a significantly lower alpha-delta ratio than those with missense variant in *STXBP1* at all ages past infancy (**Fig. 3**). Previous work has demonstrated electrographic differences in different genes within the same disorder.[4] In a novel analysis, we find significant electrographic differences between variant classes within the same gene.

Nevertheless, after infancy we find that three individuals with a missense variant have elevated alpha-delta ratios closer to those seen in individuals with a PTV at a similar age. One of these individuals has three elevated alpha-delta ratio recordings from ages 1.86-2.23 years. Notably, 10 days after this last recording they came to the emergency department due episodes of pain and feeding intolerance which resolved after gastrostomy tube placement. After this, their alpha-delta ratio dropped as seen in an EEG recorded 14 days later and in all their subsequent 17 available EEG recordings. While this could be coincidental, it could also indicate a significant adverse deterioration in their disease presentation resulting in the large drop in their alpha-delta ratio. If the latter is true, it provides further evidence of alpha-delta ratio as a biomarker for severity in *STXBP1*-related epilepsy disorders. Unfortunately, the other two individuals with elevated alpha-delta ratios have only one or two recordings available after infancy.

There are four data points from two individuals after infancy with a PTV that display an alpha-delta ratio below 0.1. Three of these originate from the same individual with a deletion of exon 4 of the gene. When raising the threshold to an alpha-delta ratio of less than 0.15 there are 8 data points from individuals with a PTV, six of which arise from this same individual. This individual, however, appears to resemble what is typically expected of an individual with a PTV in *STXBP1*.[5,6] They have an onset of infantile spasms at 5 months of age, global developmental delay, hypotonia, and feeding difficulty. At 10 years of age, they are still almost nonverbal, speaking only several words at a time which are lost and gained periodically. Moreover, their seizures have been consistently well-controlled with medication.

#### Seizure frequency prediction

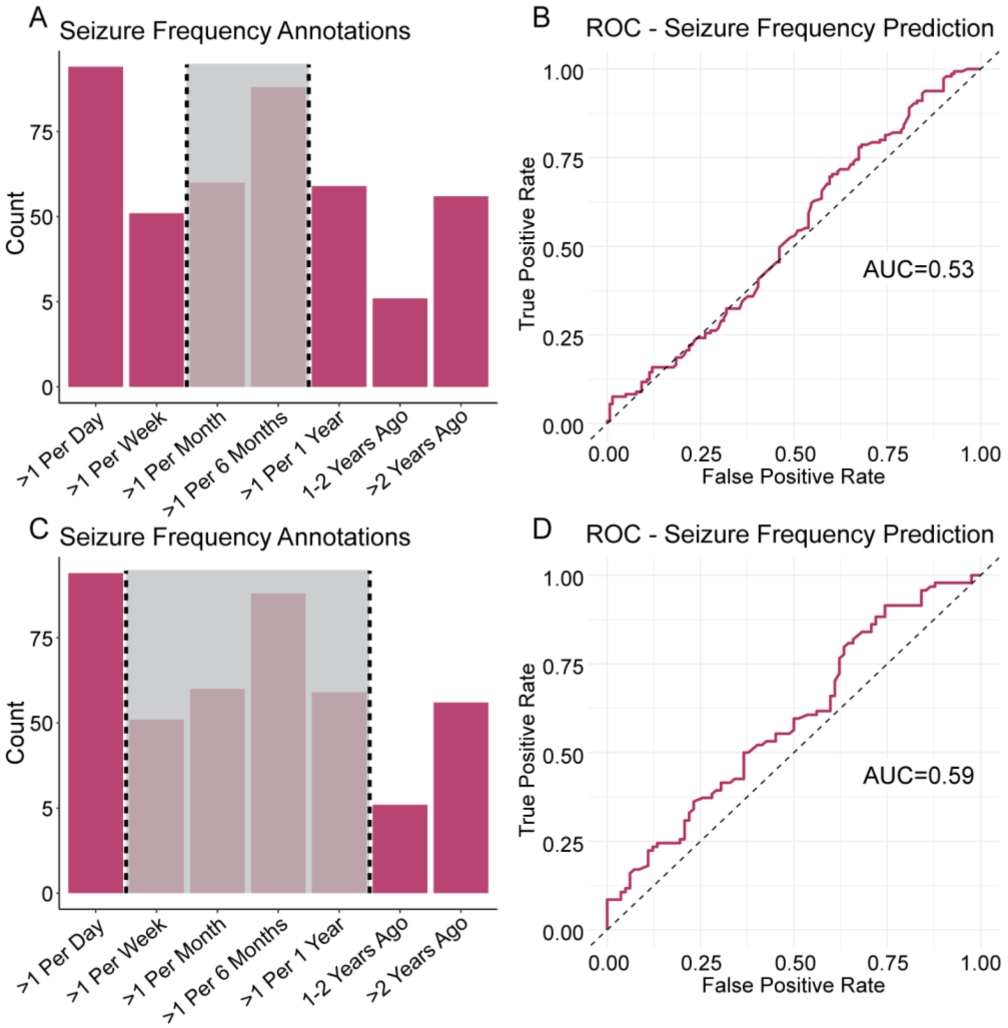


**Supplementary Figure 1 Seizure frequency prediction models with differing binary cutoffs.** The distributions and their binary cutoffs are displaying in (**A**) and (**C**). Grey shaded boxes indicate the seizure frequency annotations that were removed. The left and right side of the dashed vertical lines indicate the remaining dataset, each side indicating a separate binary outcome of 'severe' and 'moderate' seizure frequency, respectively. The performance of the random forest models from the respective datasets are displayed in the ROC curves in (**B**) and (**D**).

In our seizure frequency prediction models, we converted clinically annotated seizure frequencies to a binary moderate vs poor seizure control. We split annotations by '>2 years ago', '1-2 years ago', '>1 per 1 year', '>1 per 6 months', and '>1 per month' for moderate seizure control and '>1 per day' and '>1 per week' for poor seizure control (**Fig. 5A**). Our main analyses demonstrated that spectral features provided no correlates of seizure control (**Fig. 5B**). To ensure that this was not a consequence of our binary cut-offs, we tried several other combinations. All other model methods and characteristics remained constant, including use of age-matching and leave-one-out cross validation.

We first tried removing categories of seizure control that lay between the extremes of either binary category. In our broader genetic epilepsy group, removing ‘>1 per 6 months’ and ‘>1 per month’ resulted in an AUC of 0.53. In individuals with a genetic diagnosis, this resulted in an AUC of 0.60. Removing ‘>1 per week’, ‘>1 per 6 months’, ‘>1 per month, and ‘>1 per 1 year’ resulting in a slightly higher AUC of 0.59 in the broader genetic epilepsy cohort. In individuals with a genetic diagnosis, this resulted in an AUC of 0.61. In all cases, models with just age as a feature were tested as well and resulted in AUCs close to 0.50.

### Supplementary Figures

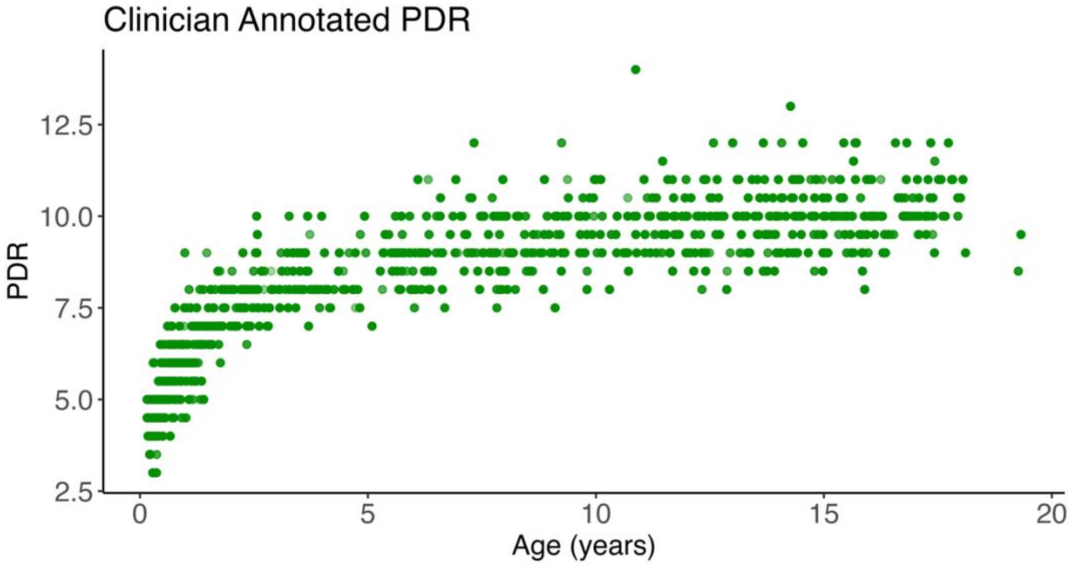


**Supplementary Figure 2 Posterior dominant rhythm across development.** Clinician annotated posterior dominant rhythm (PDR) from clinician annotations of the control cohort.

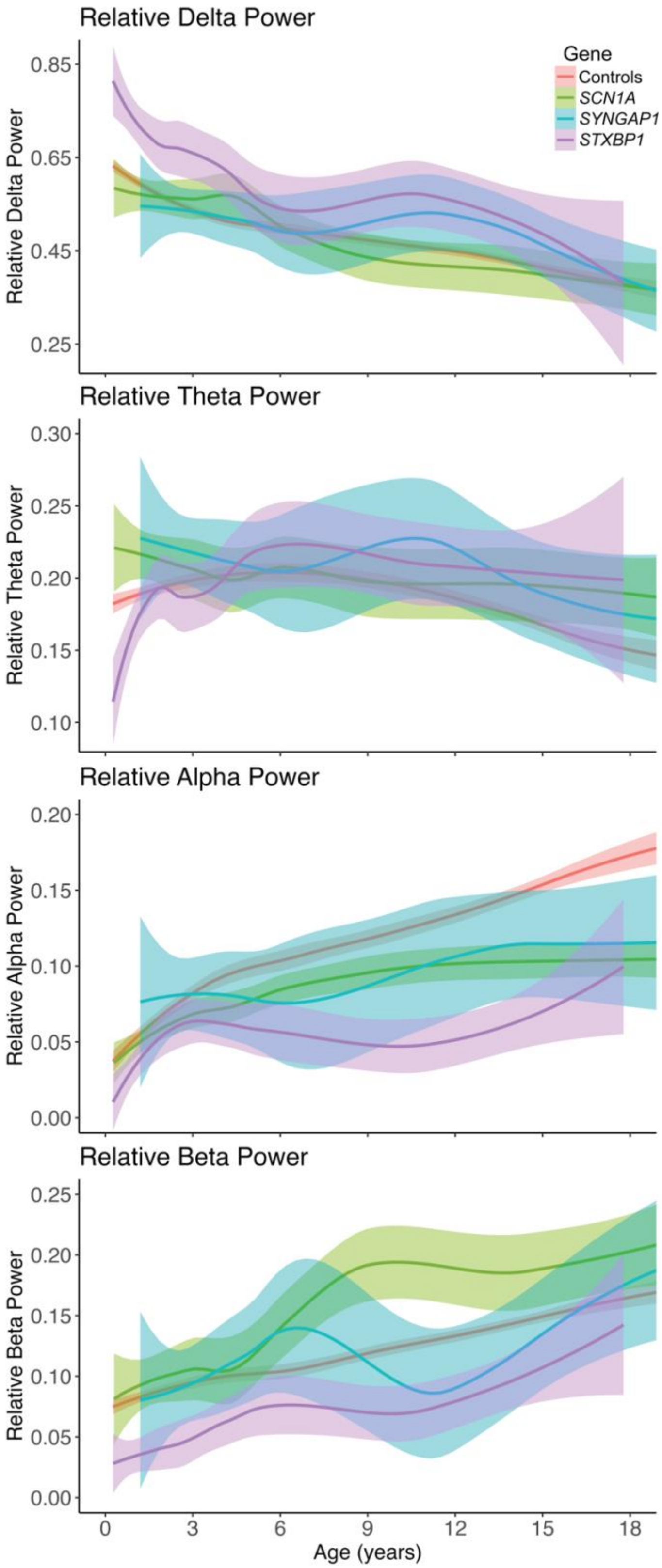


**Supplementary Figure 3 Relative bandpowers across childhood in Controls, *STXBP1*, *SYNGAP1*, and *SCN1A*.** Lines are fitted via locally weighted scatterplot smoothing to approximate a confidence interval. Displaying data from recordings from 0.25-18 years of age.

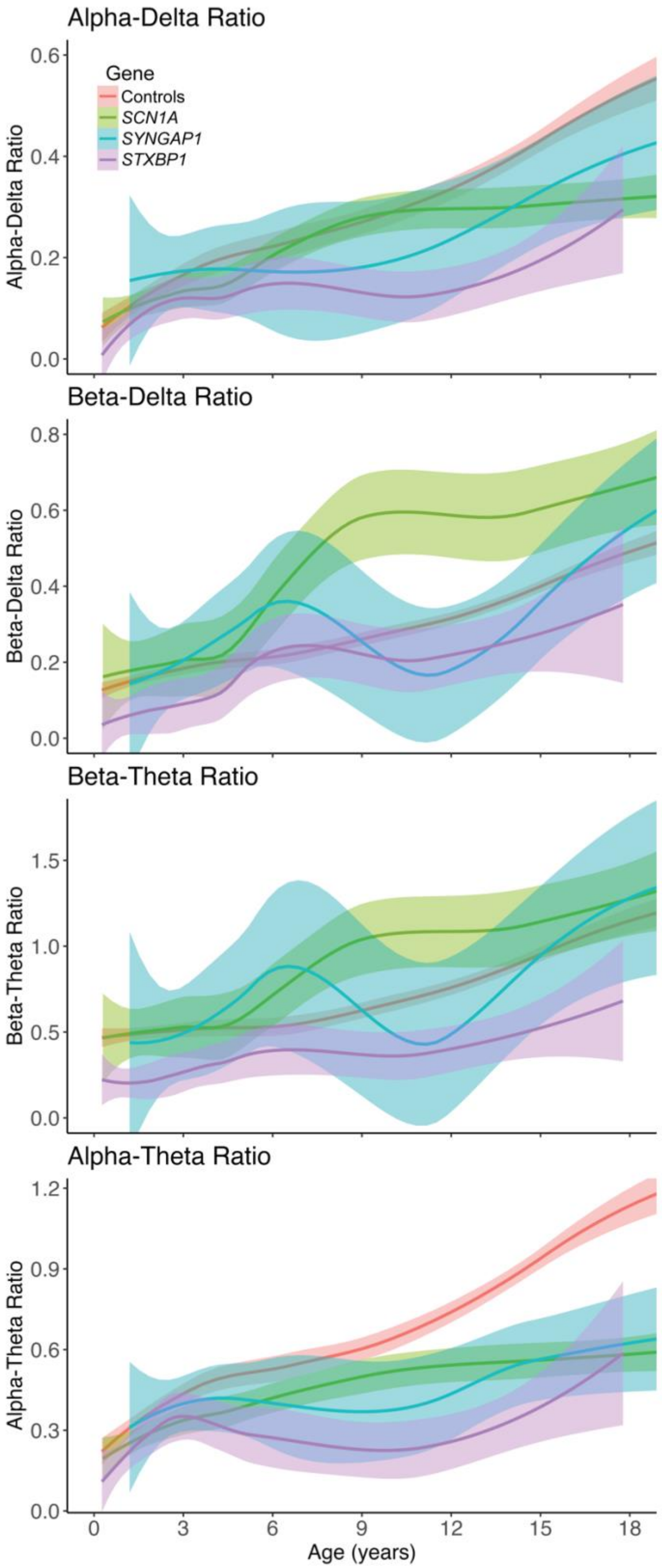


**Supplementary Figure 4 Bandpowers ratios across childhood in Controls, *STXBP1*, *SYNGAP1*, and *SCN1A*.** Lines are fitted via locally weighted scatterplot smoothing to approximate a confidence interval. Displaying data from recordings from 0.25-18 years of age.

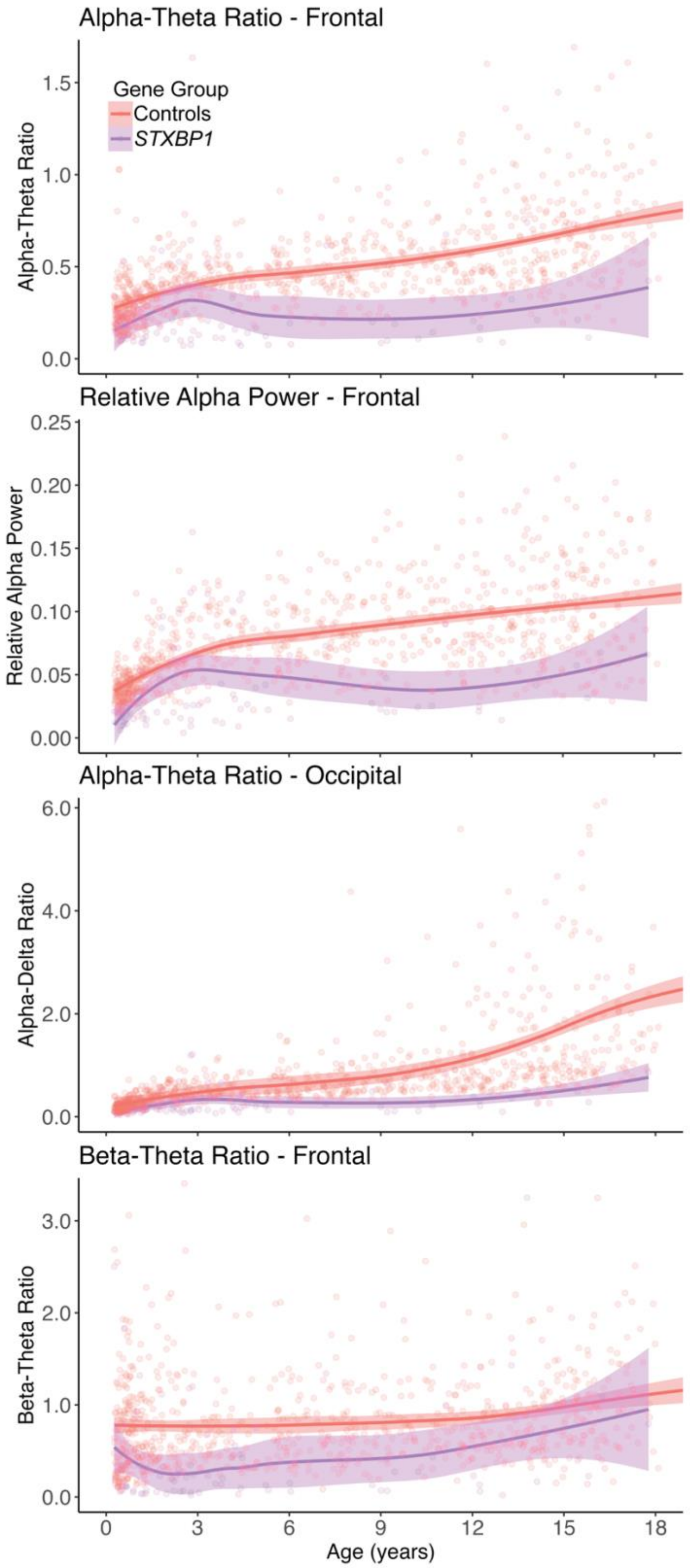


**Supplementary Figure 5 Top four features in *STXBP1* random forest models.** Displaying data from recordings 0.25-18 years of age. Some control data points are out of the set *y*-axis limits. Axis ranges were set to aid in visualization without affecting the smoothing function.

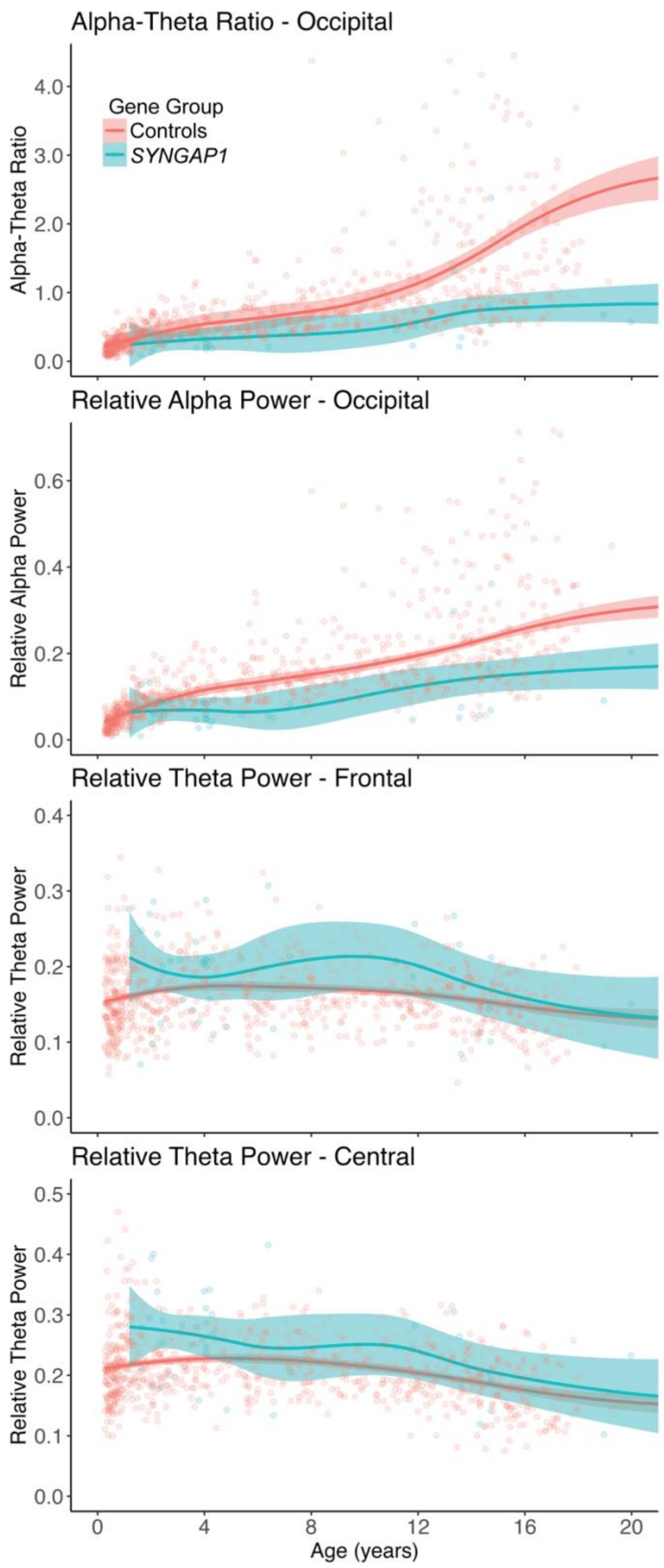


**Supplementary Figure 6 Top four features in *SYNGAP1* random forest models.** Displaying data from recordings 0.25-20 years of age. Some control data points are out of the set *y*-axis limits. Axis ranges were set to aid in visualization without affecting the smoothing function.

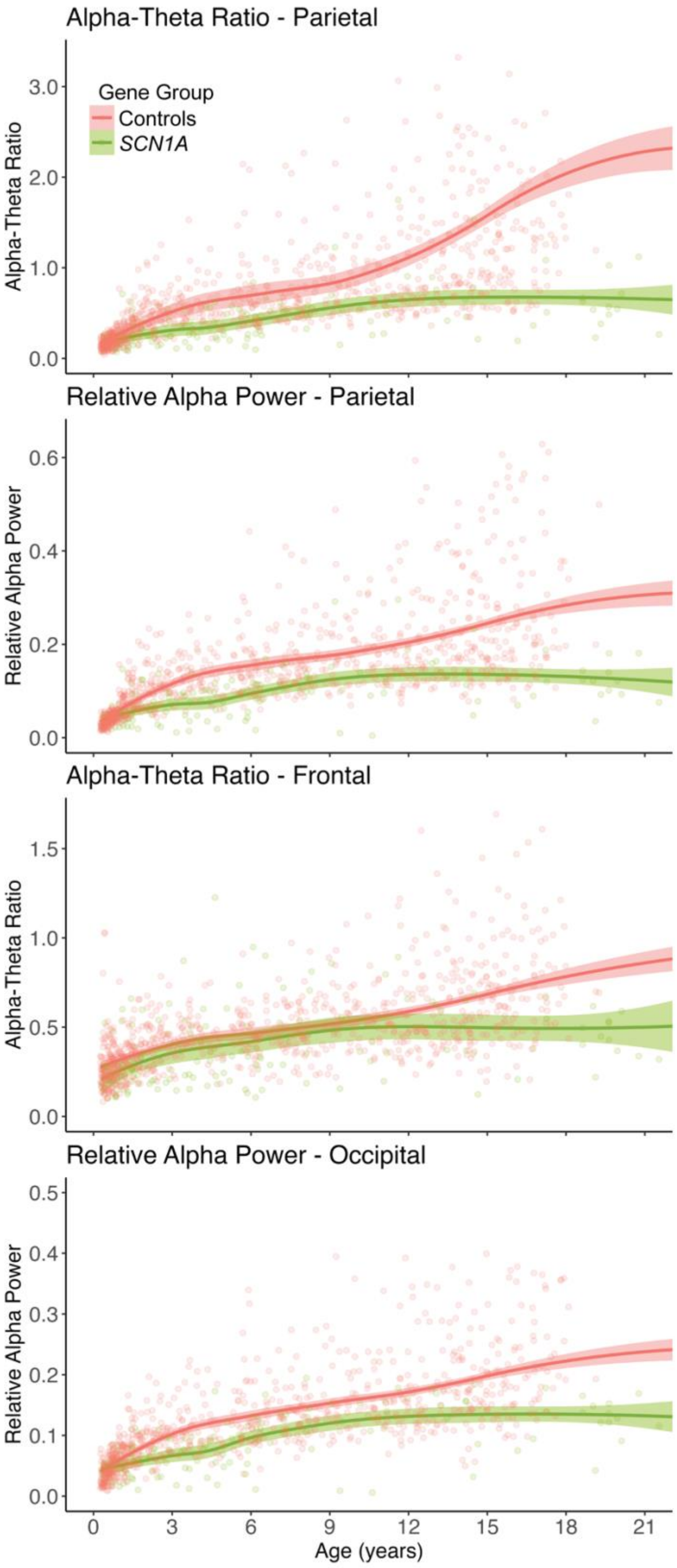


**Supplementary Figure 7 Top four features in *SCN1A* random forest models.** Displaying data from recordings 0.25-21 years of age. Some control data points are out of the set *y*-axis limits. Axis ranges were set to aid in visualization without affecting the smoothing function.

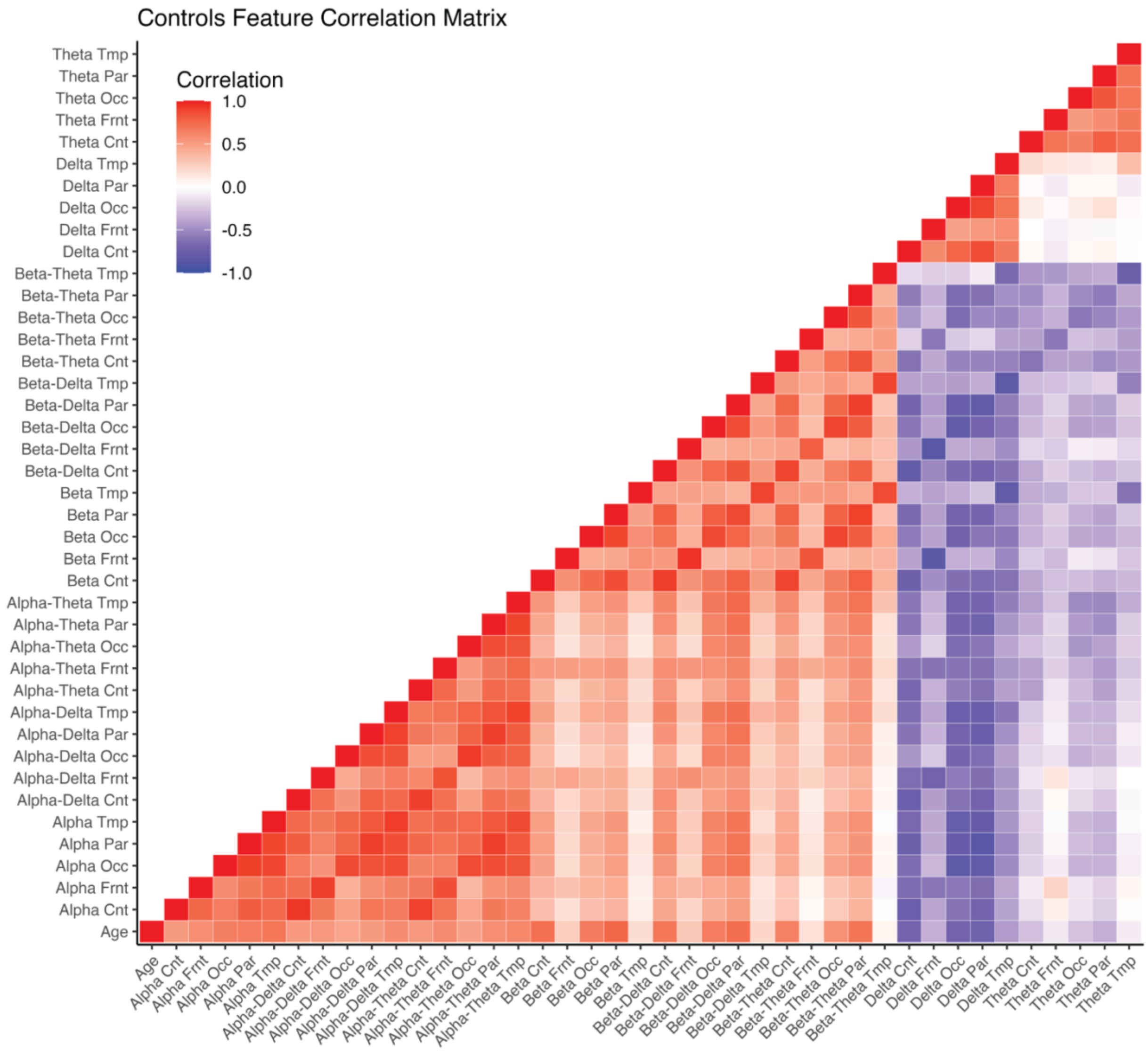


**Supplementary Figure 8 Spectral feature correlations in control cohort.** Filtered for individuals >0.25 years of age. Displayed features are all those used in the primary random forest models using spatial spectral features and age.

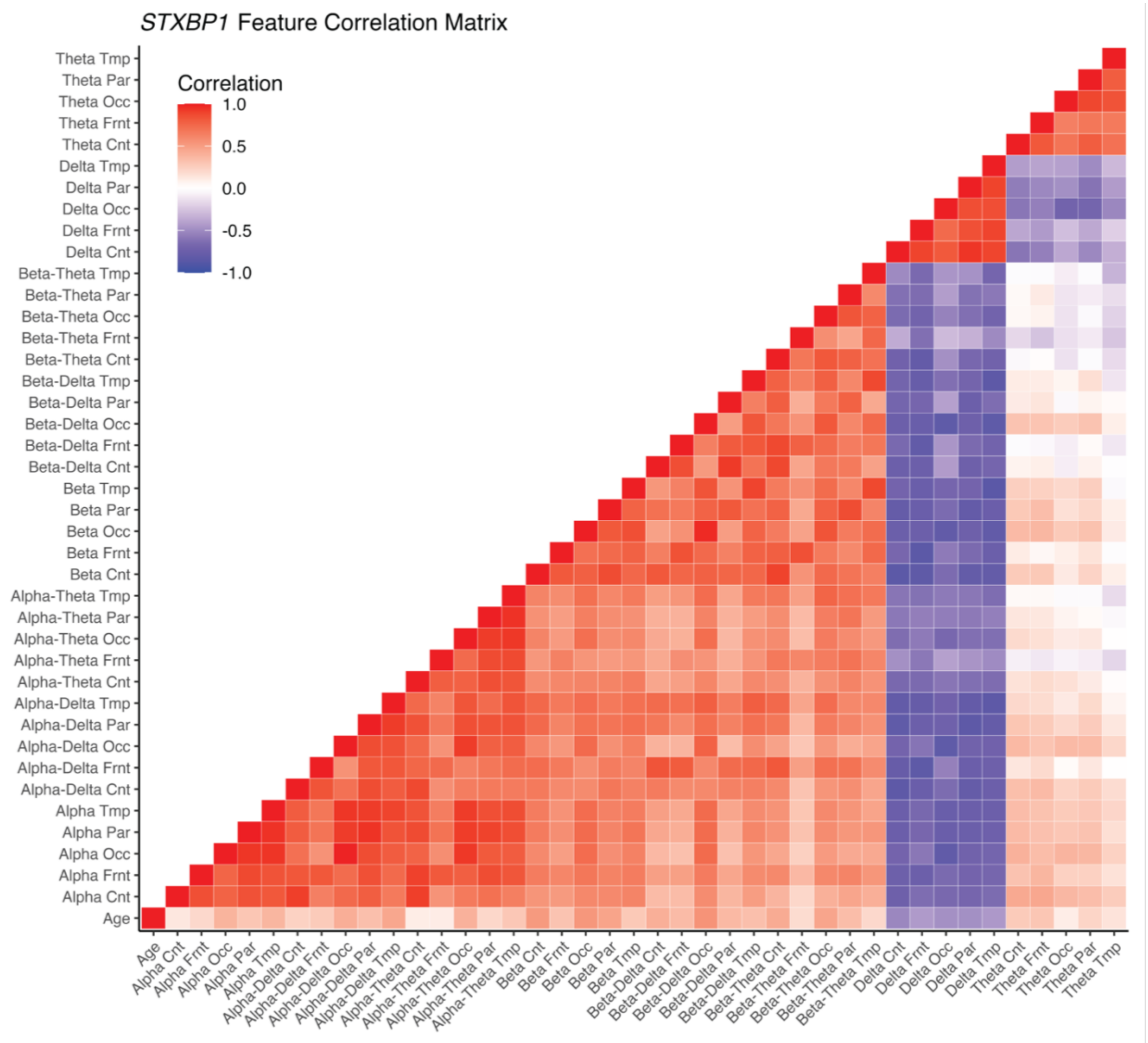


**Supplementary Figure 9 Spectral feature correlations in *STXBP1* cohort.** Filtered for individuals >0.25 years of age. Displayed features are all those used in the primary random forest models using spatial spectral features and age.

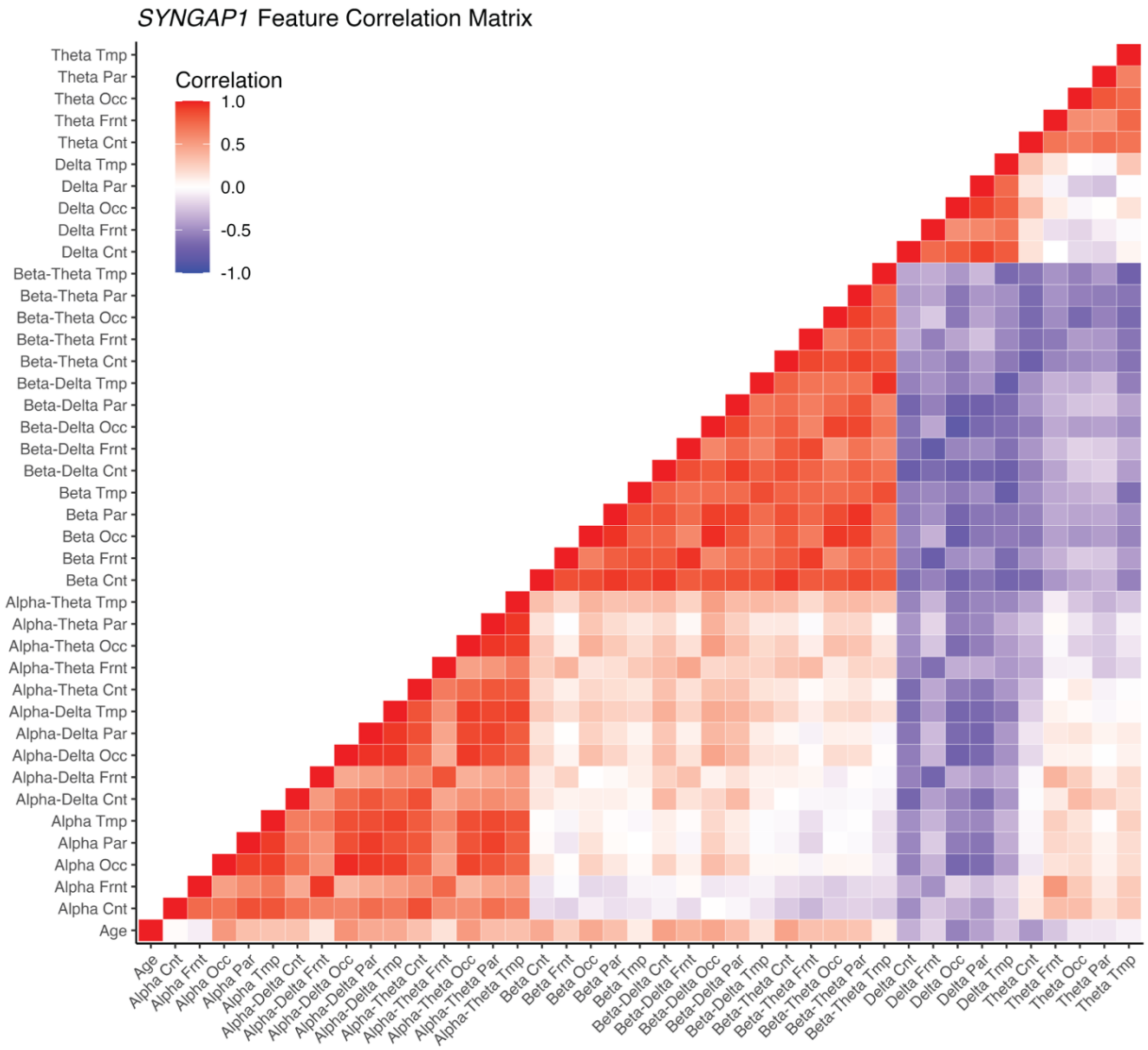


**Supplementary Figure 10 Spectral feature correlations in *SYNGAP1* cohort.** Filtered for individuals >0.25 years of age. Displayed features are all those used in the primary random forest models using spatial spectral features and age.

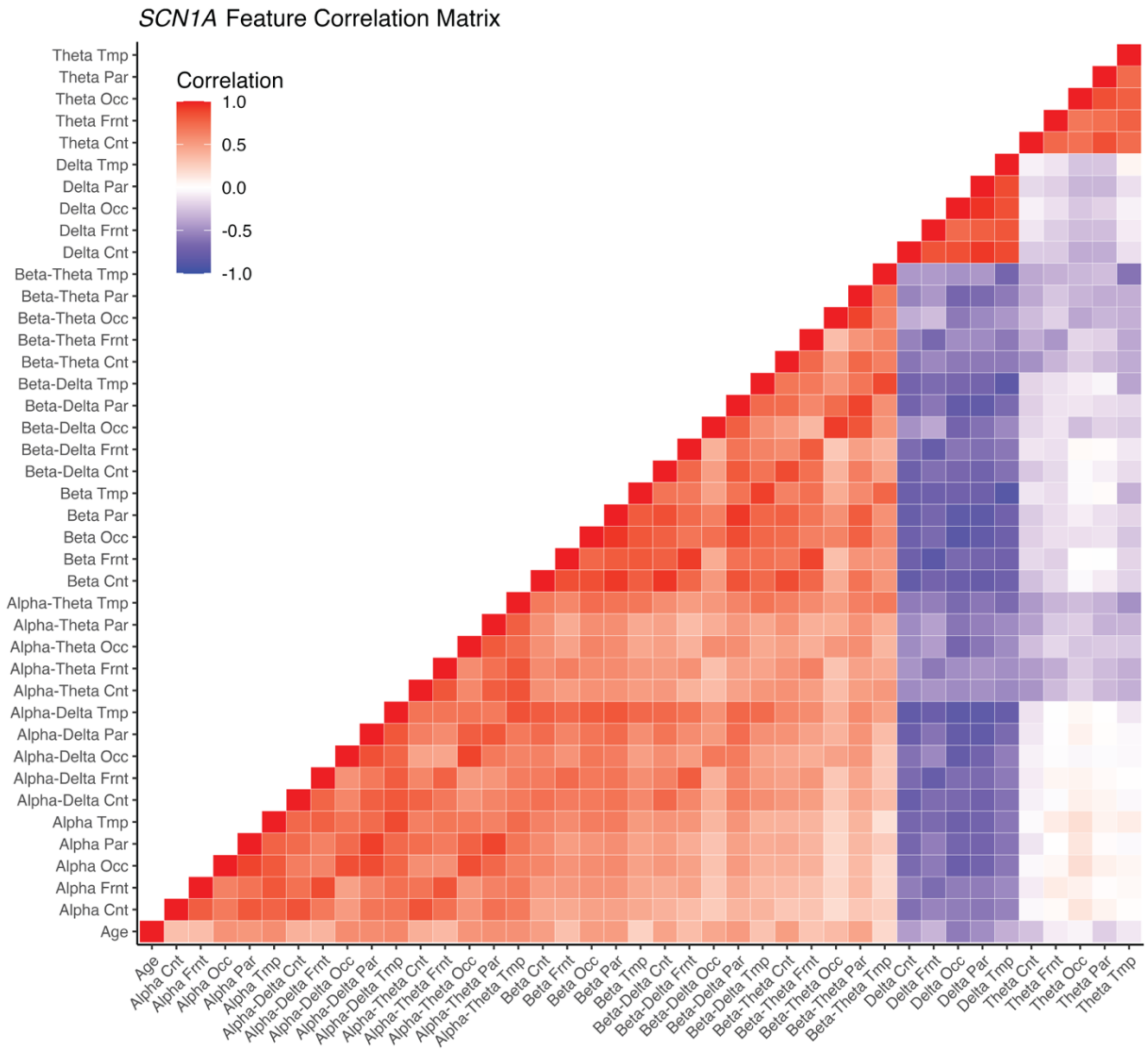


**Supplementary Figure 11 Spectral feature correlations in *SCN1A* cohort.** Filtered for individuals >0.25 years of age. Displayed features are all those used in the primary random forest models using spatial spectral features and age.

### Supplemental Tables

| | Controls | *STXBP1* | *SYNGAP1* | *SCN1A* | Seizure Frequency | GMFM |
|---|---|---|---|---|---|---|
| Standard EEG (<60 min) | 725 | 61 | 30 | 101 | 314 | 306 |
| EEG Monitoring (4 hour in Lab) | 6 | 9 | 0 | 6 | 5 | 8 |
| Ambulatory EEG Monitoring (Home) | 170 | 10 | 7 | 35 | 98 | 61 |
| Longterm monitoring EEG with Video (inpatient) | 29 | 15 | 9 | 12 | 23 | 25 |

*EEGs were grouped into broader categories

**Supplementary Table 2 Number of each EEG recording type in each cohort.**

| Age Range of EEG & GMFM (years) | Maximum Absolute Age Difference (years) |
|---|---|
| < 1 | 0.0833 |
| 1-1.5 | 0.17 |
| 1.5-2.5 | 0.25 |
| 2.5-3.5 | 0.33 |
| 3.5-4.5 | 0.50 |
| 4.5-5.5 | 0.67 |
| 5.5-6.5 | 0.75 |
| 6.5-7.5 | 0.917 |
| 7.5-8.5 | 1.083 |
| 8.5-9.5 | 1.25 |
| 9.5-12 | 2 |
| 12-17 | 3 |

**Supplementary Table 3 Filter criteria for GMFM and EEG pairing.** First column indicates range of ages in which the EEG and GMFM were recorded. Second column indicates the threshold for the absolute difference of age of recording between the EEG and GMFM acceptable for that age range. If multiple EEGs match a single GMFM, the GMFM-EEG pair closest in age was taken for analysis.
